# Blood-based Genomic and Cellular Determinants of Response to Neoadjuvant PD-1 Blockade in Patients with Non-Small-Cell Lung Cancer

**DOI:** 10.1101/2021.12.07.21267340

**Authors:** Xi Zhang, Rui Chen, Wenqing Li, Shengchao Zhang, Mengju Jiang, Guodong Su, Yuru Liu, Yu Cai, Wuhao Huang, Yuyan Xiong, Shengguang Wang

## Abstract

**Background:** Despite the improved survival observed in PD-1/PD-L1 blockade therapy, there still is a lack of response to the anti-PD1 therapy for a large proportion of cancer patients across multiple indications, including non-small cell lung cancer (NSCLC)

**Methods:** Transcriptomic profiling was performed on 57 whole blood samples from 31 NSCLC patients and 5 healthy donors, including both responders and non-responders received anti-PD-1 Tislelizumab plus chemotherapy, to characterize differentially expressed genes (DEGs), signature pathways, and immune cell subsets regulated during treatment. Mutations of oncogenic drivers were identified and associated with therapeutic outcomes in a validation cohort with 1661 cancer patients. These multi-level biomarkers were validated and compared across different methods, external datasets and multiple computational tools.

**Results:** NSCLC patients examined and achieved pathological complete response (pCR) were considered as responders or non-responders otherwise. Expression of hundreds DEGs (FDR p<0.05, fold change<-2 or >2) was changed in blood during neoadjuvant anti-PD-1 treatment, as well as in lung cancer tissue as compared to normal samples. Enriched PD-1-mediated pathways and elevated cell abundances of CD8 T cells and regulatory T cells were exclusively observed in responder blood samples. In an independent validation cohort of 1661 pan-cancer patients, a panel of 4 top ranked genetic alterations (PTCH1, DNMT3A, PTPRS, JAK2) identified from responders in discovery cohort were found positively associated with the overall survival (p<0.05).

**Conclusion:** These findings suggest peripheral blood-based biomarkers and cell subsets could be utilized to define the response to neoadjuvant PD-1 blockade in NSCLC patients and a set of novel gene mutations is strongly associated with the therapeutic outcome of cancer immunotherapy.

## Introduction

Given the significant benefit checkpoint inhibitors (ICIs) immunotherapy achieved in cancer, it is emerging as a “common denominator” in the past decade [1]. To date, several anti-PD-1 antibodies have been approved as monotherapy or neoadjuvant immunotherapy in combination with chemotherapy or other drugs [2]. It is proved that anti-PD-1 agents with chemotherapy could restore anti-tumor activities of multiple immune cell subsets, leading to an increased overall survival [3]. Despite the impressive success of PD-1 blockade therapy, the clinical benefit of this treatment is always and only limited to a small subset of patients [4]. Advanced NSCLC is one of the first pioneers becoming a common therapeutic focus for therapy targeting PD-1 or its ligand PD-L1 [5, 6]. Although the combination of anti-PD-1 therapy with chemotherapy have showed more encouraging results in the up-front treatment of NSCLC [7], the overall response rate to PD-1/PD-L1 blockade treatment in NSCLC is still low. Taking anti-PD-1 antibody Pembrolizumab as example, the objective response rate for the unselected population with NSCLC was only 19% and the median overall survival was 12 months [8].

The outcome of anti-PD-1 treatment for NSCLC patents is associated with many factors, such as PD-L1 expression level, tumor mutation burden, mRNA expression of certain biomarker genes, neo-antigens, and the composition of tumor-infiltrating immune cells [9]. Many clinical studies as well as review articles suggested PD-L1 expression testing should become (if not yet) a companion diagnostic (CDx) assay for patient who is newly diagnosed NSCLC, while other biomarkers should be evaluated in the future [10, 11]. A couple of drug-specific CDx PD-L1 immunohistochemistry (IHC) assays were validated and recently approved to determine eligibility of patients with NSCLC for anti-PD-1 treatment [12, 13]. In addition to IHC assay which requires tissue collection, the demand of treatment outcome determinants identified from non-invasive blood test is increasing. Actually a number of new biomarkers predictive of the response to ICIs were investigated, including DNA mutational variant in NK cells [14], CD28 levels in tumor-infiltrating CD8-T cells [15], metabolite from on-treatment serum [16], local or peripheral immune cell clusters [17, 18], or combination of T cell properties and energy metabolism markers in blood [19]. The proposed biomarkers for a successful response to anti-PD-1/PD-L1 treatments mostly fall in two kinds, either molecular signatures determined by genetic tests (such as tumor mutational burden [20, 21], and mismatch-repair deficiency [22]) or signatures of the immune response (such as tumor-infiltrating lymphocytes [23] and peripheral blood immune cells [24]).

In the present study, a comprehensive analysis workflow was constructed to identify multi-level biomarker signatures using the transcriptomic profiles of whole blood from a discovery cohort and followed by validations in published datasets or an independent cohort. This workflow utilized well-recognized computational tools including *in silico* deconvolution approaches developed to profile tumor-infiltrating immune cells in tissue [25, 26] and validated on PBMC samples in our prior studies [27-29]. This workflow also adapted an upgraded variant-calling method to rate and report cancer-related DNA mutations called from RNA-sequencing (RNA-seq) data [30, 31]. Together, this study identified and validated gene expression biomarkers, peripheral immune cell sets and a panel of DNA mutational markers that are expected to be predictive or responding to neoadjuvant anti-PD-1 treatment in NSCLC patients.

## Result

### Responder DEGs are potential gene expression biomarkers that alter during neoadjuvant anti-PD-1 treatment

Differential expression analysis was performed on the transcriptomic profiles between all pre- and post-treatment blood samples to identify differentially expressed genes (DEGs) in response to the treatment. In total 927 protein-coding genes were identified as significant DEGs (FDR p <0.05 and Log2(fold change) >1 or <-1) in comparison of unpaired samples and 1211 DEGs were identified from paired samples (**Figure 2A, Supplementary Table S1**). Notably, most of the DEGs (876 out of 927) identified from unpaired samples are also detected in pairwise comparison although the latter one used less samples (**Figure 2A, Supplementary Figure S1A**). In terms of the comparison of paired samples, 2298 and 1025 DEGs were identified from responders and non-responders respectively while 834 Common DEGs are shared in both (**Figure 2B, Supplementary Figure S1B**). As annotated in Figures 2A and 2B, 8 top ranked genes were selected (FDR p <0.05 and Log2(fold change) >1.5 or <-1.5) from the Common DEG lists described above and their mRNA expression levels were examined by qRT-PCR for validation (**Figure 2C**). We next looked up the expression profiles of these genes in cancer tissues database (GENT2). Interestingly, 7 out of 8 gene showed consistent alteration from tumor to normal lung tissue versus pre- to post-treatment blood samples (p<0.01) (**Figure 2D**). A strong agreement to this result is observed in a larger lung cancer database (LCE) with meta-analysis across multiple independent cohorts (**Supplementary Figure S2**).

**Figure 1.**
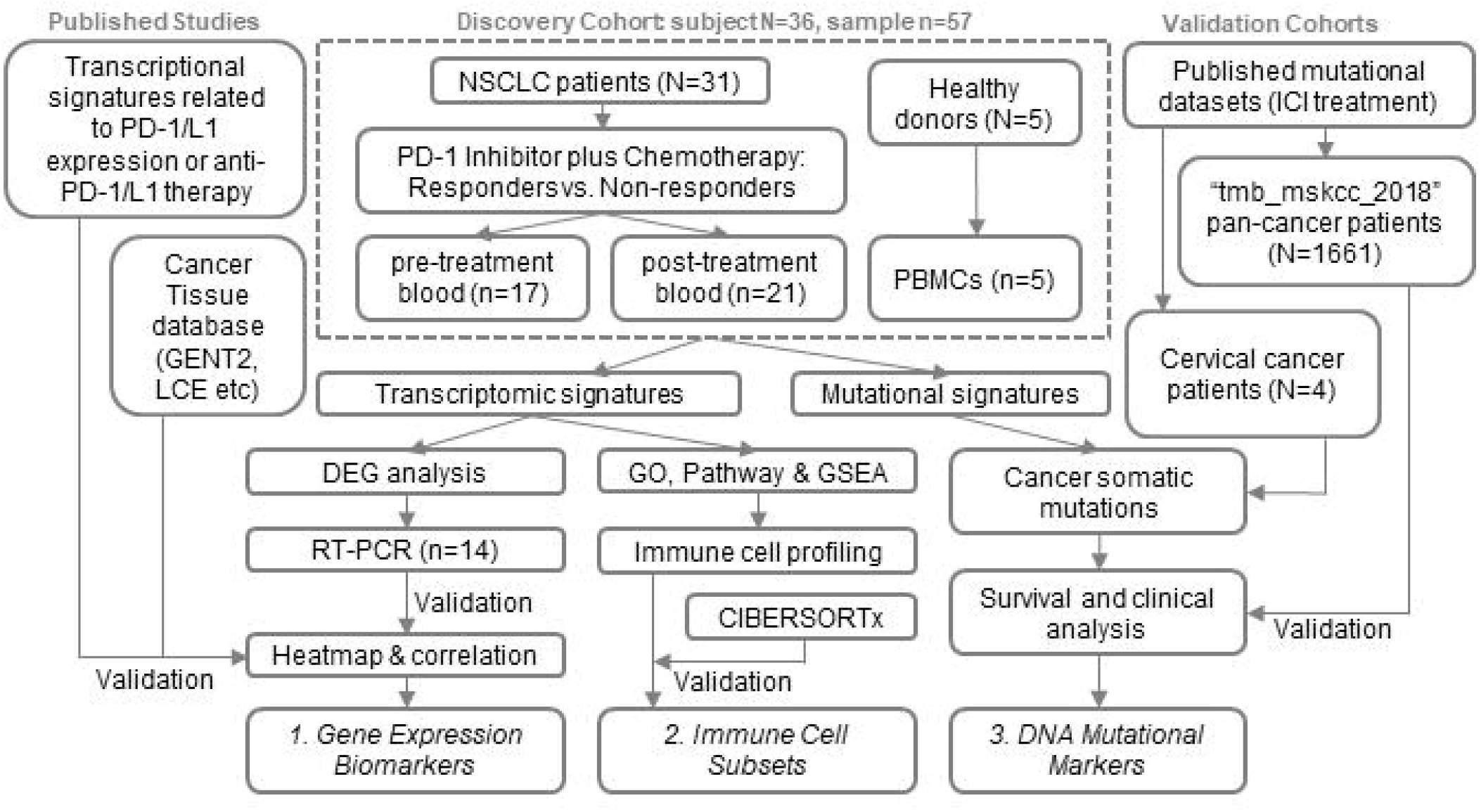
Workflow diagram of the study to identify blood-based signatures. NSCLC, non-small cell lung cancer; vs., versus; DEGs, differently expressed genes.

**Figure 2.**
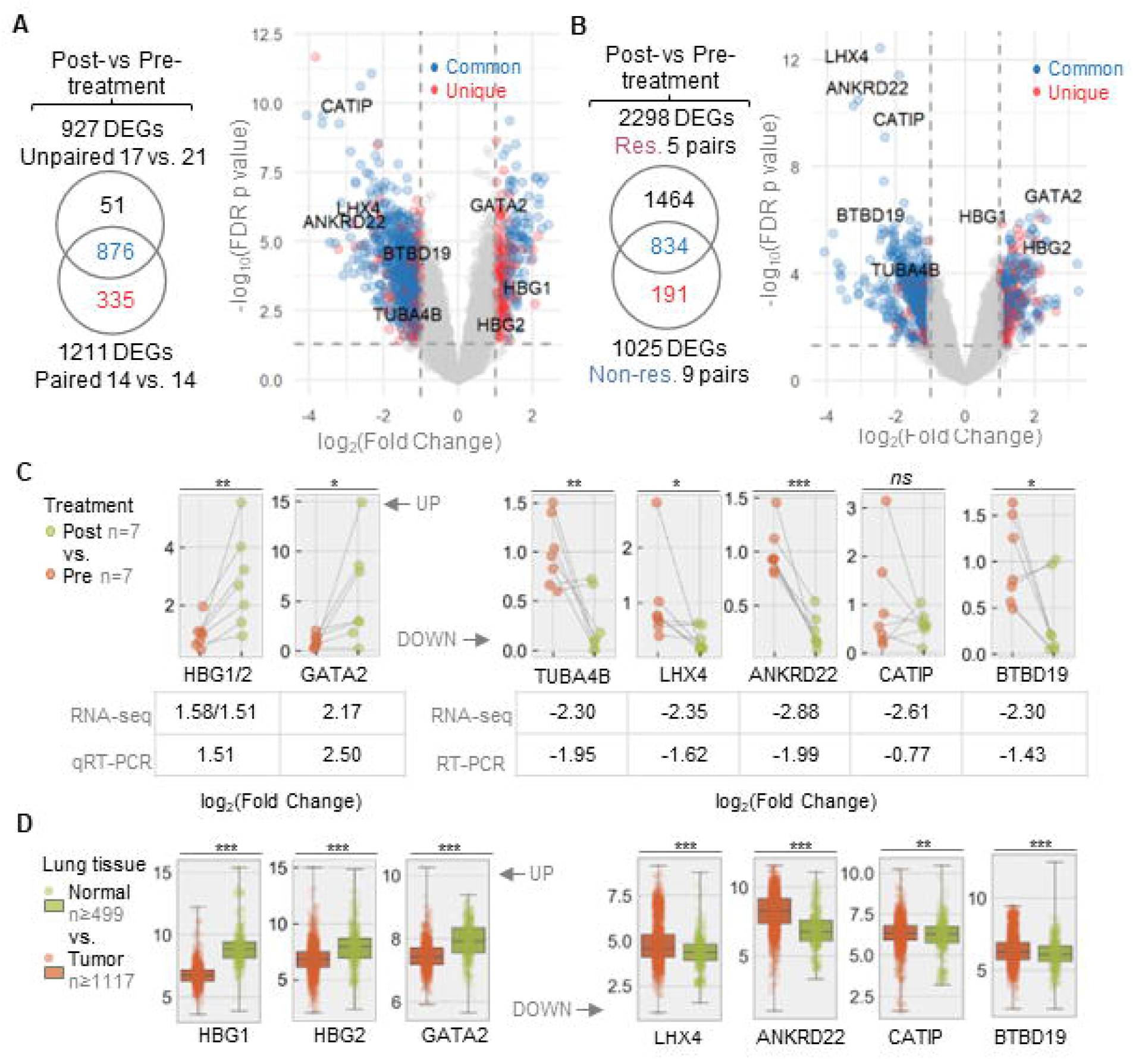
Identification and validation of transcriptomic signatures altered during neoadjuvant anti-PD-1 treatment. A) Venn diagram and volcano plot of overall differentially expressed gens (DEGs) identified in unpaired and paired comparisons of post-versus pre-treatment blood samples. Unique DEGs identified from pairwise comparison and Common DEGs identified from both pairwise and unpaired comparisons are color-coded as annotated. B) Venn diagram and volcano plot of DEGs identified in comparison of post-versus pre-treatment blood samples from non-responders. DEGs only seen in non-responders (Unique) and DEGs seen in both non-responders and responders (Common) are highlighted. The 8 Common DEGs with their names annotated in Figures 2A and 2B are genes used for qRT-PCR validation. C) Relative mRNA expressions of 8 Common DEGs validated by qRT-PCR and compared to RNA-seq results. The mean fold changes identified from both methods are provided after binary logarithmic conversion (Log_2_(mean fold change)). Genes HBG1 and HBG2 were combined in qRT-PCR because they were detected by same primer set. D) Relative mRNA expression changes of same DEGs in normal lung and lung cancer tissues (data from GENT2 database). Dot plots and box plots, if shown, represent the individual values and the average values of the mRNA levels. N, normal lung tissue; T, lung cancer tissue; v, versus; *, p<0.05; **, p<0.01; ***, p<0.001.

Next, we sought to test our hypothesis that these DEGs may serve as gene expression biomarkers during neoadjuvant anti-PD-1 treatment. Firstly only the significant DEGs from responders, either exclusively from responders (Unique DEGs) or Common DEGs, showed overlay with published transcriptional signatures that either correlating with PD-L1 expression [32] or responding to anti-PD1 therapy [33] in cancer patients (**Figure 3A**). In consistent with the RT-PCR results of Common DEGs (**Figure 2C**), a cross-method validation is also seen in 5 representative genes from Unique DEGs (**Figure 3B**). Secondly, it is notable that the Common DEGs are more significantly regulated (higher fold change and lower p value) than the Unique DEGs (**Figure 3B**). However, the Common DEGs change displayed a low correlation (correlation coefficient R^2^ = 0.29) between responders and non-responders (**Figure 3C**), suggesting the immune activation boosted by anti-PD-1 may stimulate different molecular mechanisms in responders and non-responders. Lastly, the sample similarity matrix (**Figure 3D**) as well as hierarchical clustering (**Figure 3E, Supplementary Figure S3**) results were generated using Unique DEGs and Common DEGs. In both cases, only Unique DEGs organized pre-treatment samples into a distinct group from the post-treatment samples and its gene expression profile changed significantly across pre-, post-treatment and HC samples.

**Figure 3.**
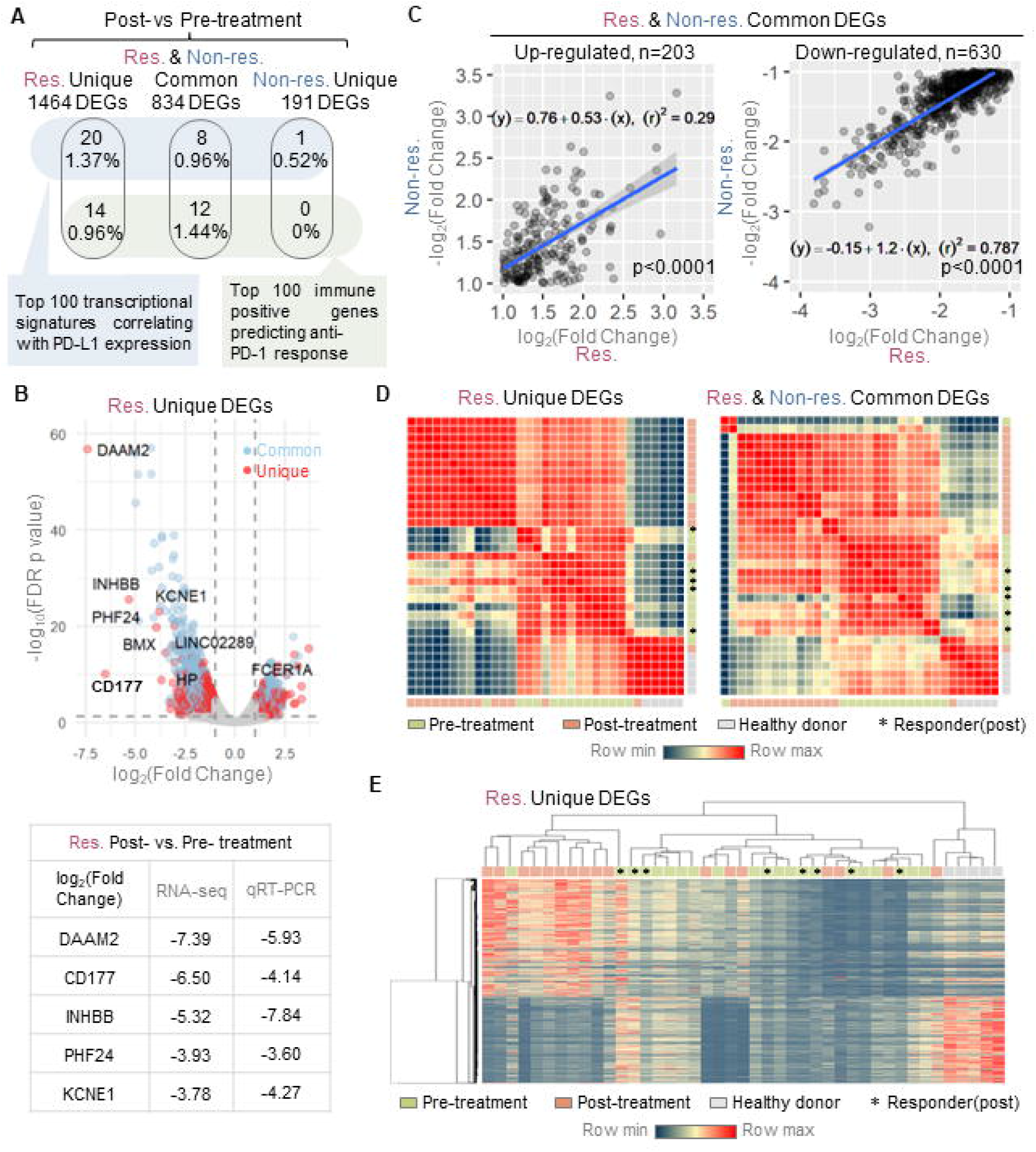
Characterization of DEGs identified from responders as gene expression biomarkers. A) Venn diagrams showing the overlay between DEG subgroups with top ranked transcriptional signatures correlating with PD-1 expression in multiple indications including NSCLC (blue box) or those responding to anti-PD-1 therapy in melanoma (green box). Unique DEGs were identified from either responders or non-responders while Common DEGs were identified from both patient groups. B) Volcano plot of DEGs identified in comparison of post-versus pre-treatment blood samples from responders. DEGs only seen in responders (Unique) and DEGs seen in both responders and non-responders (Common) are highlighted. The mean fold changes in RNA-seq and RT-PCR are provided (in the format of Log_2_(mean fold change)). C) Correlation scatter plots of the fold change (log2) of Common DEGs (left, up-regulated; right, down-regulated) between responders (n=14) and non-responders (n=14). The p value and R squared (square of the correlation coefficient) were produced by a Pearson correlation analysis. The linear regression line and its equation were generated from a simple linear regression analysis. D) Sample similarity matrices from Unique or Common DEGs to visualize the mix-ups of 14 paired patients samples and 5 healthy donor samples. E) The hierarchical clustering of all study samples according to their profiles of responder Unique DEGs. The heatmap visualized the relative expression level of each DEG. Pre- and post-treatment status, responder labels are color-coded and annotated.

### PD-1-mediated pathways and regulatory T cells are distinct signatures in responders during the treatment

We also asked which pathways and immune cell responses account for a successful response to the treatment. Unlike the striking difference observed in the numbers of DEGs identified from responders vs. non-responders (**Figure 2B**), the pathways and GOs identified from these two groups are quite comparable. Not only the total numbers are close but also a majority of them are commonly seen in two groups, at least in KEGG pathways (211 out of 246 and 237) and GOs (2144 out of 2634 and 2728) (**Figure 4A, Supplementary Figure S4 & S5**). Uniquely, the result in Reactome database showed about one quarter (215 out of 817) of the total pathways (**Figure 4A**) and 12 out of the top 20 pathways are exclusive in responders (**Figure 4B**) and non-responders (**Figure 4C**). Interestingly, several key pathways related with PD-1 regulatory network are overrepresented and only overrepresented in responders, including signaling of PD-1, CD3, TCR, CD28, IL-1 and IFN-γ. Gene set enrichment analysis (GSEA) analyses were performed across all available collections and the top ranked gene sets were compared in distinct-directional classes (up and down regulations) (**Supplementary Figure S6**). All GSEA results suggested substantial changes in immune cell subsets such as a decrease in monocytes compared to plasmacytoid DCs and an increase in CD4+ T cells compared to myeloid cells (**Figure 5A & 5B, Supplementary Figure S7**).

**Figure 4.**
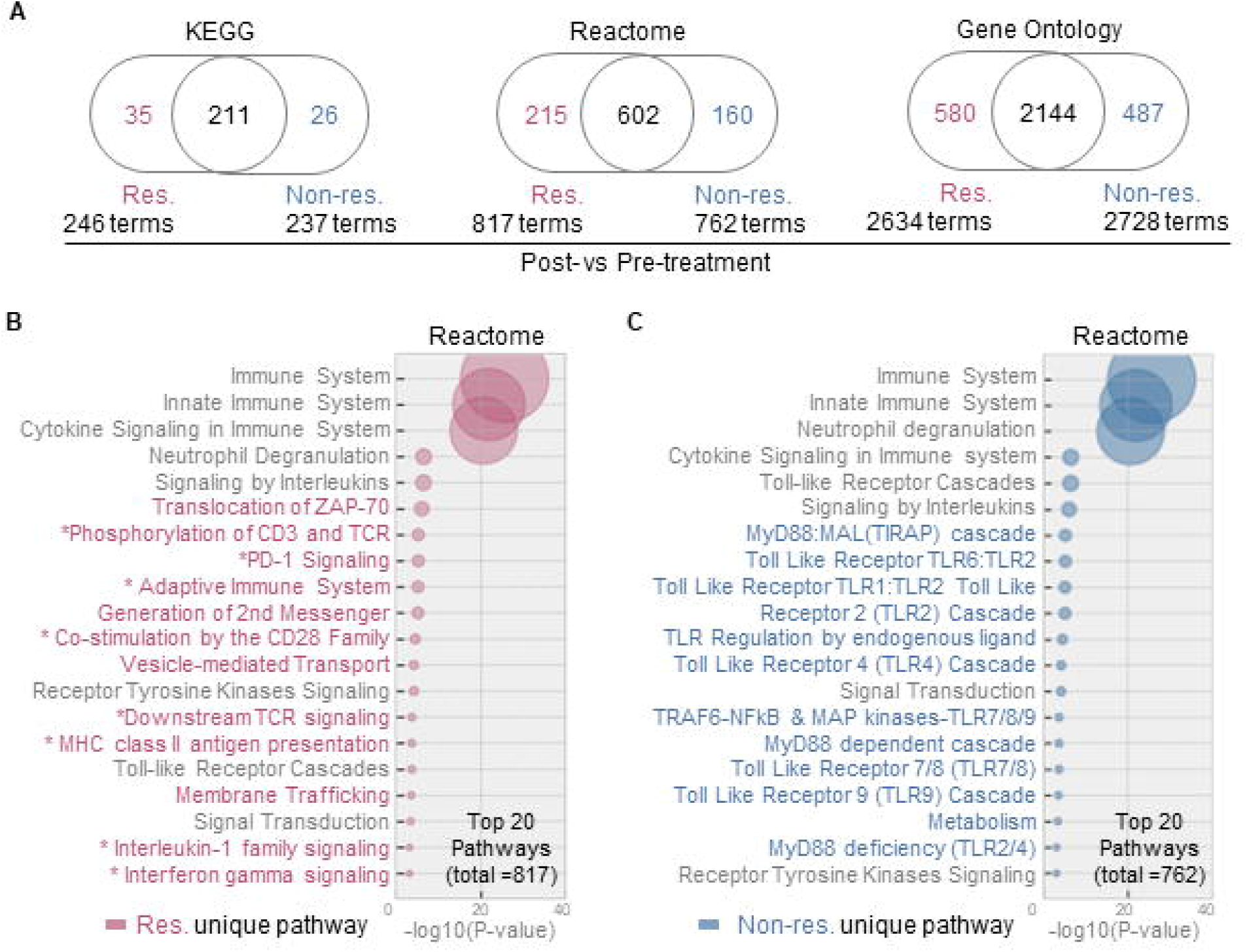
Analysis of pathways and GOs that specifically regulated in responders during the treatment. A) Venn diagram visualization of the significant KEGG pathways, Reactome pathways and gene ontology (GO) items regulated after treatment. The numbers of terms regulated in responders, non-responders and both are provided respectively. B & C) Bubble plots of the top 20 pathways regulated in responders (B) and non-responders (C). Bubble with bigger size stands for smaller p value and higher significance. Names of unique terms are colored in red (responder) or blue (non-responder) while the shared terms are annotated in grey text. Res, responders; Non-res, non-responders.

**Figure 5.**
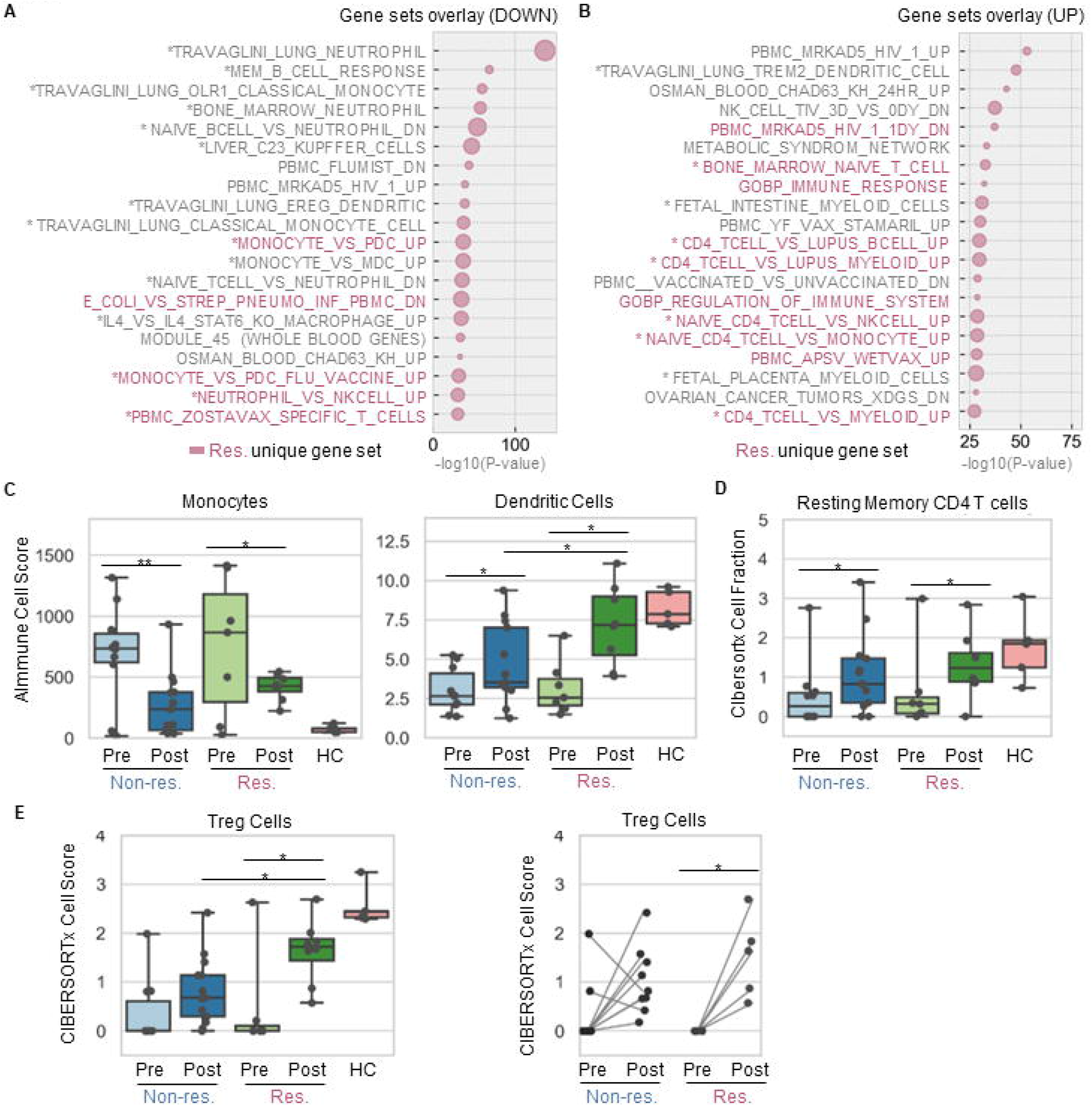
GSEA and immune cell abundance profiling to identify key immune cell subsets responding to the treatment. A & B) Bubble plots of the top 20 gene sets down-regulated (A) and up-regulated (B) in responders. Bubble with bigger size stands for smaller p value and higher significance. Names of unique terms are colored in red (responder) while the shared terms are annotated in grey text. Bubble with bigger size stands for higher k/K value ratio and larger fraction of gene was matched with a certain reference gene set. C & D) Cell abundance scores as calculated by AImmune (C) and CIBERSORTx (D) of immune cell subsets are significantly changed in both responders and non-responders. E) Cell abundance scores of regulatory T cells (by CIBERSORTx) is only significantly changed in responders but not in non-responders. Dot plots and box plots show individual values and average value of the scores. Connecting lines indicate the pairwise relationship between pre- and post-treatment samples. Res, responders; Non-res, non-responders; HC, healthy control. *, p<0.05; **, p<0.01. All p values were calculated for pairwise comparisons.

The immune cell abundance change was further elucidated by *in silico* leukocyte deconvolution approach adopted from our previous work (AImmune) [27-29] as well as a classic machine learning tool CIBERSORTx [26] (**Supplementary Figure S8 & S9**). In agreement with the GSEA results, a significant composition decrease in monocytes and a sturdy increase of DCs were observed in AImmune (**Figure 5C, Supplementary Figure S8**) while a substantial increase of resting memory CD4+ and CD8+ T lymphocytes were demonstrated in CIBERSORTx (**Figure 5D, Supplementary Figure S9**). While the changes of the cell subsets above were actually seen in both responders and non-responders, the abundance of macrophages, CD8 T cells and regulatory T cells were elevated and only elevated in responders (**Supplementary Figure S8, Figure 5E**).

### A panel of mutational markers was validated through an independent cohort to predict sensitivity to PD-1 blockade

The Cancer-Related Analysis of Variants Toolkit (CRAVAT) was applied in the RNA-seq data from our discovery cohort (**Figure 1**) in order to identify somatic mutations that predict survival benefit for anti-PD-1 treatment (**Supplementary Figure S10**). Top 50 mutated genes (classifier gene-level p values <0.05 and FDR = 0.05, ranked by CHASM score) were identified from responders, non-responders and healthy donors, with minority of the genes are shared by two or more groups (**Figure 6A**). The scores generated from CHASM (driver classifier) and VEST (pathogenicity classifier) are all closed to 1, suggesting these mutations are confidently classified as cancer drivers and likely are pathogenic (**Figure 6B**). Three out of all 39 mutational markers (TNFAIP3, BRCA1, ASXL1) identified exclusively from non-responders in our NSCLC cohort were shared with the top 50 mutational markers from progressive disease patients of an independent cervical cancer cohort (GEO repository: GSE205247) (**Figure 6C**). It is worth noting 3 shared genes here are highly ranked as the top 1st, 3rd and 4^th^ mutations in the discovery cohort (bold in **Figure 6B**).

**Figure 6.**
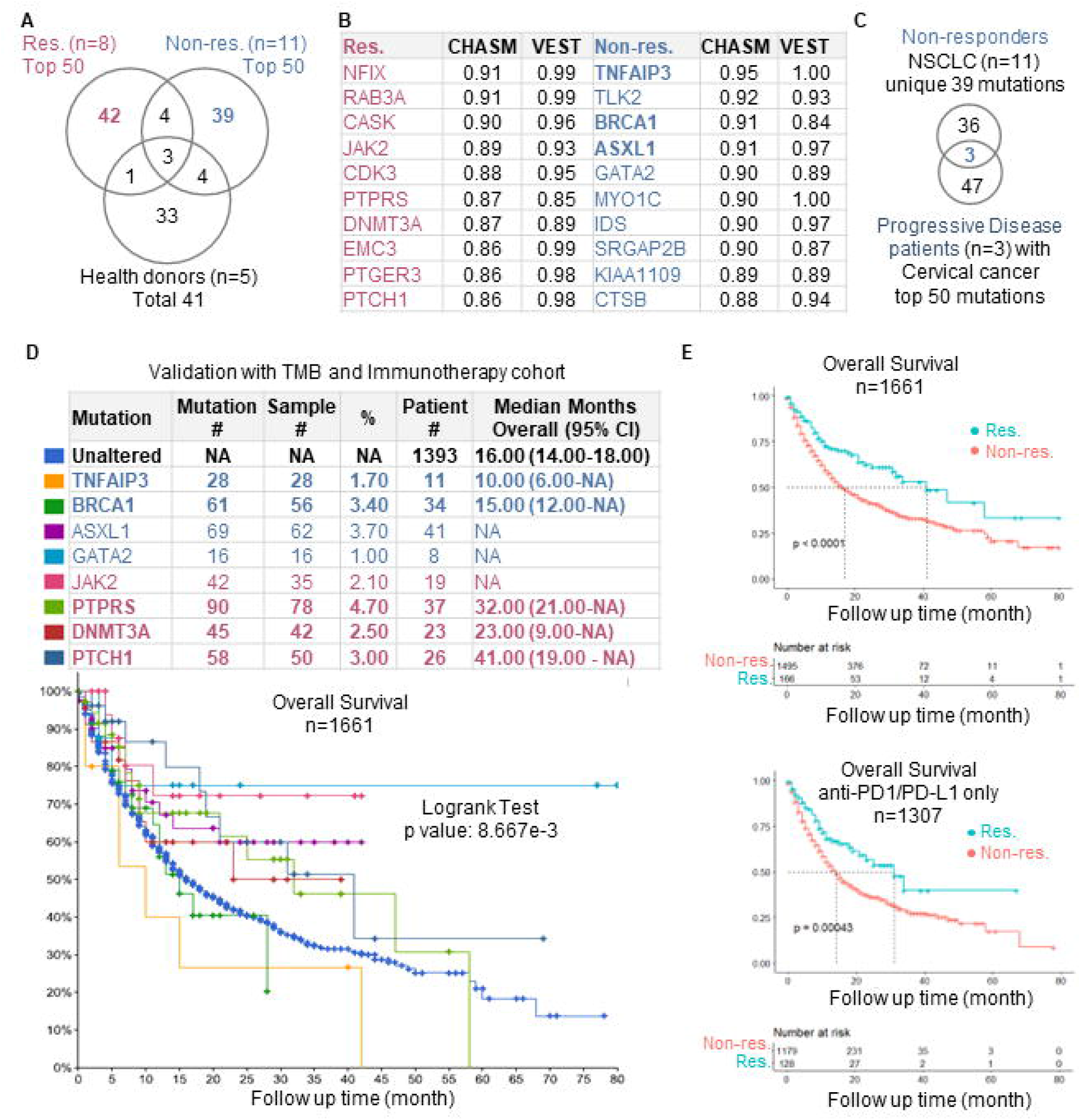
Identification and validation of blood-derived mutational markers to neoadjuvant anti-PD-1 treatment. A) Venn diagram showing the overlay between the top 50 or all gene-level mutations identified from blood samples in responders, non-responders and healthy donors. B) Top 10 mutated genes exclusively identified from responders and non-responders and ranked by CHASM and VEST scores. C) Venn diagram comparing the mutated genes from non-responders of NSCLC patients and mutated genes from progression disease patients with cervical cancer (published dataset). The 3 overlaid genes are bold in table showed in B). D) Mutation status of the mutated genes in an independent pan-cancer cohort (TMB and Immunotherapy cohort, n=1661) and the Kaplan Meier overall survival curves of pan-cancer patients grouped by 8 mutational markers individually. E) The Kaplan Meier overall survival curves of two patient groups defined by 4 responder markers combined as one panel. Curves were generated for all immunotherapy patients (upper) and patients received anti-PD1/PD-L1 therapy only. P value was generated from by Log-Rank test which compares the survival distributions in independent groups.

In another much larger validation cohort (“tmb_mskcc_2018”) with 1661 pan-cancer patients [34], 8 out of all 20 top-ranked gene mutations from the discovery cohort were detected patients who all received PD-1 or CTLA-4 blockade treatment (**Figure 6D**). The Kaplan-Meier curves illustrates the significantly difference of overall survival (p<0.01) across 9 patient groups as defined by 8 mutational markers. The overall median survival among patients with no mutation is 16 months, while the median survivals (if available) are shorter (10-15 months) in patients with non-responder mutations and much longer (23-41 months) in patients with responder mutations (**Figure 6D**). Moreover, the mean survival was consistently extended (41 vs 17 months in Res. vs Non-res. for all patients, 31 vs 14 months in Res. vs Non-res. for anti-PD1/PD-L1 treated patients) in a patient group defined by a combination of all 4 genetic alterations (PTCH1, DNMT3A, PTPRS, JAK2) identified from responder (**Figure 6E**). No significant difference was observed in patient groups defined by a combination of non-responder mutations (**Supplementary Figure S11**).

## Discussion

Given the majority of tumor patients fail respond to anti-PD-1/PD-L1 therapy [35], a lot of effort made by clinical trials led to the identification of predictive biomarkers to select treatment responders and the discovery of underlying mechanisms during treatment for improving current treatment modalities [36, 37]. Although PD-1/PD-L1 IHC expression test has been widely evaluated as a predictive biomarker, as an invasive tissue test it is still not implemented as a routine diagnostic test for anti–PD-1/PD-L1 therapy worldwide [38]. Part of the reason is lack of universal standard of antibody for detecting PD-L1 expression and no mention it is still controversial whether PD-1/PD-L1 expression is an inadequate determinant of the treatment.

The overall hypothesis of the present study is the blood-based molecular signatures that differently expressed in blood samples from responders versus non-responders, and thus provide additive prognostic value in patient selection. In order to test this hypothesis, the present study employed an integrated workflow and identified multi-level response biomarkers from a discovery cohort, including gene expression biomarker, immune cell subsets and DNA mutational markers. The discovering results were then further validated by different methods (RNA-seq and qRT-PCR), multiple databases (GENT and LCE), innovative tools (AImmune and CIBERSORT) as well as independent cohort (tmb_mskcc_2018 etc.) By comparing responders and non-responders during the treatment, our data provided new evidences suggesting key driving genes, targetable pathways, peripheral immune cells, and most importantly mutational determinants which are detectable by blood test and of predictive value towards the therapy outcome and clinical efficacy.

At the beginning of this study, a group of Common DEGs that changed during the treatment were found consistently altered in overall comparisons of both responders and non-responders (**Figure 2A & 2B**). These genes were ranked as top DEGs in an earlier version of the discovery cohort and thus were selected for RT-PCR validation and database investigation. Our finding of these genes was significant as them were, on one hand, and regulated by similar folds between RNA-seq and qRT-PCR (**Figure 2C**); on the other hand, their expression levels were also changed from tumor to normal tissue sample (**Figure 2D & Supplementary Figure S2**). This signature group contains a hematopoietic transcription regulator (GATA2) [39], a metabolism mediator (ANKRD22) [40, 41], and a transcription factor involved in differentiation control (LHX4) [42], which is consistent with the current understandings that immunological and metabolism mechanisms are enriched in post-treatment and responder patients [43]. Moreover, several of them were proven to regulate the oncogenesis of lung cancer or respiratory diseases. For instance, ANKRD22 as we found down-regulated in NSCLC patients after therapy, was reported to promote the progression of NSCLC by enhancing cell proliferation [44]. The expression of HBG1/2 is induced in normal lung tissue after smoking and is downregulated again when tumors form in smokers [45]. More importantly, a recent report also determined HBG1 was consistently upregulated in blood samples of patients with stage III/IV NSCLC from the 2^nd^ anti-PD-1 antibody treatment through to the 5^th^ treatment [46]. This observation seems dependent on stage and histology as both protein and mRNA levels of HBG1/2 were found down-regulated in stage I NSCLC adenocarcinomas [47]. Because these gene were idented from non-responders as well, these findings suggested the anti-PD-1 treatment may change transcription levels in cancer-relevant genes that not necessarily indicate a positive response.

More importantly, the present study identified Unique DEGs of responders which together with the Common DEGs showed exclusive overlays with two sets of published signature genes that are related with PD-L1 expression or anti-PD-1 therapy response. The first set of genes are the top 100 (out of the total 1325) transcriptional correlates of PD-L1 expression [32]. The second set of genes were the 100 immune-positive genes used for predicting melanoma patient response to anti-PD-1 therapies as identified by integrating MHC I-association prediction with the TCGA transcriptomic data from pre- or on-treatment skin cutaneous melanoma samples (anti-PD-1 or combination of anti-PD-1 and anti-CTLA-4) [33]. The overlays between these published signature genes with newly identified DEGs from responders (**Figure 3A)** are not high (likely due to differences between study subjects, cancer types and sample types) but significantly greater than the DEGs only in non-responders. This result clearly proposed the biomarker value in responder DEGs rather than non-responder DEGs, which was further confirmed by sample similarity and hierarchical clustering (**Figure 3D & 3E)**. The predictive values of these newly identified markers for immunotherapy are either novel or only recently reported in other cancer indications. The top 1^st^ ranked DEGs identified in responders in this study is DAAM2, which was recently proved to predict high sensitivity to chemotherapy, anti-EGFR therapy and immunotherapy for in pancreatic cancer patients [48]. The other DEG PHF24 as we identified down-regulated in responders here were found highly co-expressed with gene LAMP2 which confers prognostic value to indicate worse survival of esophageal cancer patients [49].

Given PD-1/PD-L1 pathway is a central mediator of immunosuppression, PD-1 pathway blockade therapy is known to crosstalk and fine-tune with T cell activation and differentiation as well as other immune cell functions. When analyzing the Reactome pathways and GOs regulated in responders, it makes a lot sense to only see PD-1 signaling and PD-1-regulated signaling cascades (such as CD3, TCR, CD28, IL-1 and IFN-γ) in responders but not in non-responders (**Figure 4B)**. In contrast, PD-L1 expression and PD-1 checkpoint pathway in cancer is a KEGG pathway regulated in both responders and non-responders, although the significance is much weaker in the latter one (**Supplementary Figure S4**). Our finding is supported by several studies either concluded the expression of PD-1 or PD-L1 might not be qualified as a consistent biomarker for NSCLC patients received combination of chemotherapy plus immunotherapy [50], or questioned the practice of dichotomizing the range of PD-L1 expression levels is adequate for patient stratification [51]. Our GSEA analysis and immune cell abundance analysis (**Figure 5**) further sustained our findings in pathway and GO analyses, which are also backed up by a broad range of published studies. Firstly, since monocyte-to-DC differentiation attenuates CD8+ T cell response and other T-cell populations expanding [52], this differentiation was found significantly higher during successful PD-1 blockade therapy [53-55]. In line with that, our results observed DCs and macrophages were amplified in responders while mononuclear phagocytes as their precursor cells responded oppositely. Secondly, CD4 T cell memory was another cell subset we found expended during the treatment. Notably a longitudinal study recently reported that higher percentages of effector, CD62Llow CD4+ T cells, was detected in peripheral blood of responder patients with non–small lung cancer [56]. It was proposed that the abundance of certain CD4+ T cell subsets in peripheral blood before treatments is an effective predictor of responses in NSCLC patients [57]. At last, regulatory T cell (Treg cell) was found significantly elevated only in responders here, suggesting a specific role of Treg cell during PD-1 mediated immune suppression that may determine the outcome of anti-PD-1 therapy. PD-1 blockade may re-activate both CD8+ T cells and Treg cells but our finding confirmed that an effective therapy requires a skewed balance in favor of the effector CD8+ T cells rather than Treg cells [58, 59]. Summarizing, our data above proposed several novel DEGs and immune cell subsets that may serve as monitoring markers in peripheral blood of NSCLC patients. Given the fact these markers are, at least partially, independent of the PD-L1 expression levels in tissue, they are expected to bring additional values in guiding clinical decision-making.

In the last part of this study, several cancer driver genes were found significantly mutated in discovery cohort and led to extended (responder mutations) or reduced (non-responder mutations) survivals in validation cohorts (**Figure 6**). Individual diver gene mutations were previously demonstrated to promote (such as KRAS [60], TP53 [61], PTCH1 [62]) or weaken (such as JAK1/2[63], EGFR[64], PTEN [65]) the response to immune checkpoint inhibitor therapy in cancer patients. On one hand, the role of somatic mutation is consistent across cancer indications. An typical example is TNFAIP3 mutation indicates low responses to PD-1 inhibitor in NSCLC and cervical cancer patients as showed in the present study, which is also supported by a study on melanoma [66]. Another example is that the association of PTCH1 mutation with improved outcome of PD-1 blockade was seen in both colorectal cancer [62] and NSCLC (the present study). On the other hand, the role of a somatic mutation might vary across different cancer types. For example, JAK2 is associated with acquired resistance in melanoma [63] while in this NSCLC study it marks a better PD-1 blockade response. The overall impact of genome-wide mutation is to increase therapeutic sensitivity to immunotherapy as large-scale clinical trials revealed that high tumor mutational burden (TMB) level is associated with T-cell–mediated response and thus correlates with improved survivals to PD-1 plus CTLA-4 blockade in NSCLC patients [21, 67]. In line with this finding, our study highlights four newly identified mutations (PTCH1, DNMT3A, PTPRS, JAK2) as one panel is robustly predictive of clinical benefit across multiple cancer types with CTLA4 or PD-1/PD-L1 blockade therapy (**Figure 6E**).

It is important to point out the cell abundance analysis tool AImmune employed here is developed in-house and there is still room for improvement. Although this tool underwent robust testing and validation in our previous studies [27-29], the current version is only designed to cover up to 10 immune cell subsets from peripheral blood. This coverage limit is partially due to its nature as AImmune was trained and developed to profile circulating blood samples while other computational immune profiling tools including CIBERSORTx [26, 68] and TIMER [69] are often employed to enumerate tumor-infiltrating cells in tissue-dependent microenvironments. Further development and fine-tuning of AImmune tool are required and it is expected to observe more effector T cell subsets detected in responders, such as CD62Llow CD4+ T cells, CD4 Th2 cells and Treg cells [56, 57, 70]. Other limitations of this study include its relatively small sample size, which decreased statistical power and flexibility in drawing statistical conclusion. Given the high molecular heterogeneity among different NSCLC subtypes, the number of patients sequenced in this study might be too small to provide strong statistical power or drawn informative conclusions. It is encouraged that our current findings and results should be further confirmed by a larger cohort with long-term follow-up and time series analysis.

In summary, the integrated analyses employed in the current study identified blood-based molecular and cellular signatures that are associated with the responsiveness and overall survival of PD-1/PD-L1 blockade therapy plus chemotherapy.

## Method

### Study cohort and overall workflow

In order to characterize blood-based determinants of positive response to PD-1 blockade treatment, this study built a discovery cohort collecting 57 whole blood samples from 31 patients visited Tianjin Cancer Hospital from 2020 to 2021 and 5 healthy donors (**Figure 1**). Five blood samples were collected from healthy donors and included as control samples in the discovery cohort. All patients were diagnosed with non-small cell lung cancer (**Table 1**) and underwent treatment of anti–PD-1 monoclonal antibody Tislelizumab [71, 72], in combination with standard chemotherapy according to the histology. The majority of the patients were male (86.7%), over 60-year-old (66.7%), ever-smokers (80%), and classified as lung squamous cell carcinoma (SqCC) (76.7%) (**Table 1**). The protocol was approved by the local Ethics Committee and the Institutional Review Board of Norwest University (approval number: 200402001) and all subjects provided written informed consent.

**Table 1.** General characteristics of NSCLC patients in this study (N=31)

| **Characteristics** | **Count (percentage)** |
| --- | --- |
| **Gender** |  |
| Male | 27 (87.1%) |
| Female | 4 (12.9%) |
| **Age** |  |
| ≥60 | 20 (64.5%) |
| <60 | 11 (35.5%) |
| **Smoking history** |  |
| Yes | 25 (80.6%) |
| No | 6 (19.4%) |
| **Histology** |  |
| Squamous (SqCC) | 26 (83.9%) |
| Adenocarcinoma (ADC) | 4 (12.9%) |
| Large cell carcinoma (LCA) | 1 (3.2%) |
| **Stage** |  |
| □A | 10 (32.3%) |
| □B | 4 (12.9%) |
| □A | 1 (3.2%) |
| □B | 5 (16.1%) |
| □A | 5 (16.1%) |
| IIIB | 3 (9.7%) |
| □A | 1 (3.2%) |
| Unable to stage | 2 (6.5%) |
| **Responder status** |  |
| Responder (pCR) | 12 (38.7%) |
| Non-responder (non-pCR) | 17 (54.8%) |
| Not evaluated | 2 (6.5%) |

pCR, pathologic complete response

In this study, 12 patients who achieved pathological complete response (pCR) were considered as responders, 17 patients categorized as non-responders and 2 patients were not examined to determine their responsiveness status. Briefly, gene expression biomarkers responding to the treatment were identified, validated by RT-PCR and compared with published transcriptional signatures correlated with PD-1 expression or therapy response [32, 33] and lung cancer tissue datasets (GENT2: https://gent2.appex.kr; LCE: https://lce.biohpc.swmed.edu). Pathways, gene ontology items and gene sets altered during treatment were investigated via KOBAS-i portal (http://kobas.cbi.pku.edu.cn/) [73]. Secondly, peripheral immune cell subsets activated or inactivated by the therapy were characterized and valeted by two independent immune profiling tools (AImmune; CIBERSORTx: https://cibersortx.stanford.edu). Finally, DNA mutational markers were reported by CRAVAT (https://www.cravat.us/CRAVAT) and then compared and validated through independent validation cohorts contain cancer patients received immune checkpoint inhibitor therapy [34, 74] (Gene Expression Omnibus, GSE44227; cBioPortal, tmb_mskcc_2018).

### AImmune analysis and computational analysis

A novel *in silico* leukocyte deconvolution method, named AImmune, is a computational approach developed by integrating our established immune cell profiling [30] with published single cell RNA-seq data obtained from NSCLC blood samples (1071 qualified cells from one patient, GSE127471) and healthy blood samples (8369 and 7687 cells from two donors, 10X Genomics). Briefly, with the additional marker genes included, more than 30 candidate marker genes for each cell subsets in peripheral immune cell subsets (CD4-T cells, CD8-T cells, B cells, Monocytes, DCs, NK cells and NKT cells) were selected based on their expression patterns across immune cell subsets [68]. The pairwise similarity statistic of all cell subsets was computed (data not shown) between all pairs of the candidate marker genes within the normalized RNA-seq profiles (FPKM) from whole blood samples. Using the criteria (average Pearson correlation factor >0.60, p<0.01), 10-20 selected marker genes were identified as our final marker genes. The raw cell abundance score was calculated as the sum of the simple averages of the marker genes’ log2 expression, which allows comparison of cell composition across subject groups. This approach also tested a novel deconvolution model (unpublished) built by DNN (deep neural networks) algorithms and then trained by pseudo-bulk samples obtained by randomly subsampling of published scRNA-seq data [75]. Machine learning-based feature extraction (marker gene selection) was integrated for model optimization. Most of the computational analysis procedure was coded by common Python packages; scRNA-seq data was processed by R package scanpy; machine learning model was developed and tested with Python library Tensorflow. All computational analysis were performed and visualized using R version 3.6.1 or Python version 3.7.9.

### RNA isolation, transcriptomic profiling, and qRT-PCR

Buffycoat or PBMCs were isolated from whole blood. Total RNA was isolated using Trizol (Invitrogen, Waltham, MA, USA) and the purity and concentration were verified using a NanoDrop ND-1000 instrument (ThermoFisher Scientific, Waltham, MA, USA). The integrity of the RNA was assessed by a 2100 Bioanalyzer gel image analysis system (Agilent, Santa Clara,

CA, USA). Only RNA samples with an integrity number of 7 or higher were processed for RNA-seq. At least 100 ng mRNA per sample was submitted for library preparation.

Qualified RNA samples were then enriched and synthesized into two strand cDNA for library preparation. RNA-seq libraries were constructed using the TruSeq RNA Sample Prep Kit (Illumina, San Diego, CA, USA). The libraries from qualified RNA samples were sequenced in the 150 nt paired-end mode on an Illumina HiSeq 6000 platform at Novogene Bioinformatic Technology (Tianjin, China). After quality filtering (FastQC, quality value >5), over 30 billion clean reads were obtained in each library and then used for down-stream analysis.

PCR primers were designed top-ranked genes (10 for each comparison) as obtained by differential gene analysis (**Supplemental Table S2**). Real-time quantitative PCR was performed in real-time PCR systems (Bio-Rad, Hercules, CA, USA). The 2–ΔΔCT method for relative gene expression analysis was performed to calculate the mRNA levels of individual gene. Two or three replicates were measured for each sample.

### Bioinformatics and variant calling

All raw RNA-seq reads were filtered by R package trim_fastq to remove adapters, rRNA and low-quality reads. The QC criteria included: removing bases below Phred quality 20, containing over two “N”, or shorter than 75. The output reads were then indexed by aligner STAR and mapped to reference genome by BAM. Normalized read counts were generated and compared between groups to generate DEGs using R package DESeq2. Another R package countToFPKM was employed to produce FPKM for AImmune analysis. Genes that displayed at least two-fold difference in gene expression between comparison groups (false discovery rate [FDR]<0.05) were considered significant DEGs and carried forward in the analysis. Differentially expressed RNAs were illustrated in a volcano plot. Hierarchical clustering was performed to show the gene expression patterns and similarities among samples. Pathway and gene ontology (GO) enrichment analysis was carried out via an integrated platform KOBAS 3.0 [35]. GSEA analysis was carried out by searching the established MSigDB gene-set collections (C7). CIBERSORTx analysis was performed following the instruction from the portal (https://cibersortx.stanford.edu). Differences of mRNA levels and cell abundance scores were evaluated using independent *t*-tests or paired *t*-tests if pairwise samples were given. CHAVAT [30, 31].

Variant calling was performed using the HaplotypeCaller that plug-in in the Genome Analysis Toolkit v4.0 (GATK). First, RNA reads were aligned to the reference genome using the STAR aligner, then the MarkDuplicates was used to clean up data. The gatk BaseRecalibrator and gatk ApplyBQSR were used to adjust the mass fraction of original bases, detect the system errors in mass fraction, and reduce the false positives. We used only variants marked with PASS in the VCF file and filtered the variant calls with the VariantFiltration tool. The Cancer-Related Analysis of Variants Toolkit (CRAVAT) is a well-recognized informatics toolkit used in this study for variant calling from VCF files [30, 31]. This tool covers multi-level mutational analysis functions including mutation mapping and quality control, impact prediction and extensive annotation. Two Random Forest filters were employed in CRAVAT for predicting mutation impact, namely Cancer-Specific High-Throughput Annotation of Somatic Mutations (CHASM) and Variant Effect Scoring Tool (VEST). CHASM is a classifier that classifies if a mutation is an oncogenic driver while VEST rates if a mutation is pathogenic or benign.

A p value of <0.05 was considered statistically significant. Bioinformatics and statistics analyses were performed and visualized using R version 3.6.1 or Python version 3.7.9.

## Data Availability

All data produced in the present study are available upon reasonable request to the authors

## Author contributions

XZ designed the workflow, analyzed and illustrated the data, drafted and finalized the manuscript, and supervised all aspects of the study. RC built the data-analysis pipeline. WL processed samples and performed experiments. SZ and YC reviewed the manuscript and provided advice. MJ performed AImmune analysis. GS, YL and WH aided in sample collecting and processing. YX aided in experiment design, supervised experiment, and reviewed the manuscript. SW recruited patients, collected clinical information, supervised all clinical data analysis, and reviewed the manuscript.

### Acknowledgments

We thank all patients and donors who donated blood samples in this study.

