## Supplemental Figures for "Blood-based Genomic and Cellular Determinants of Response to Neoadjuvant PD-1 Blockade in Patients with Non-Small-Cell Lung Cancer"

### Slide 1
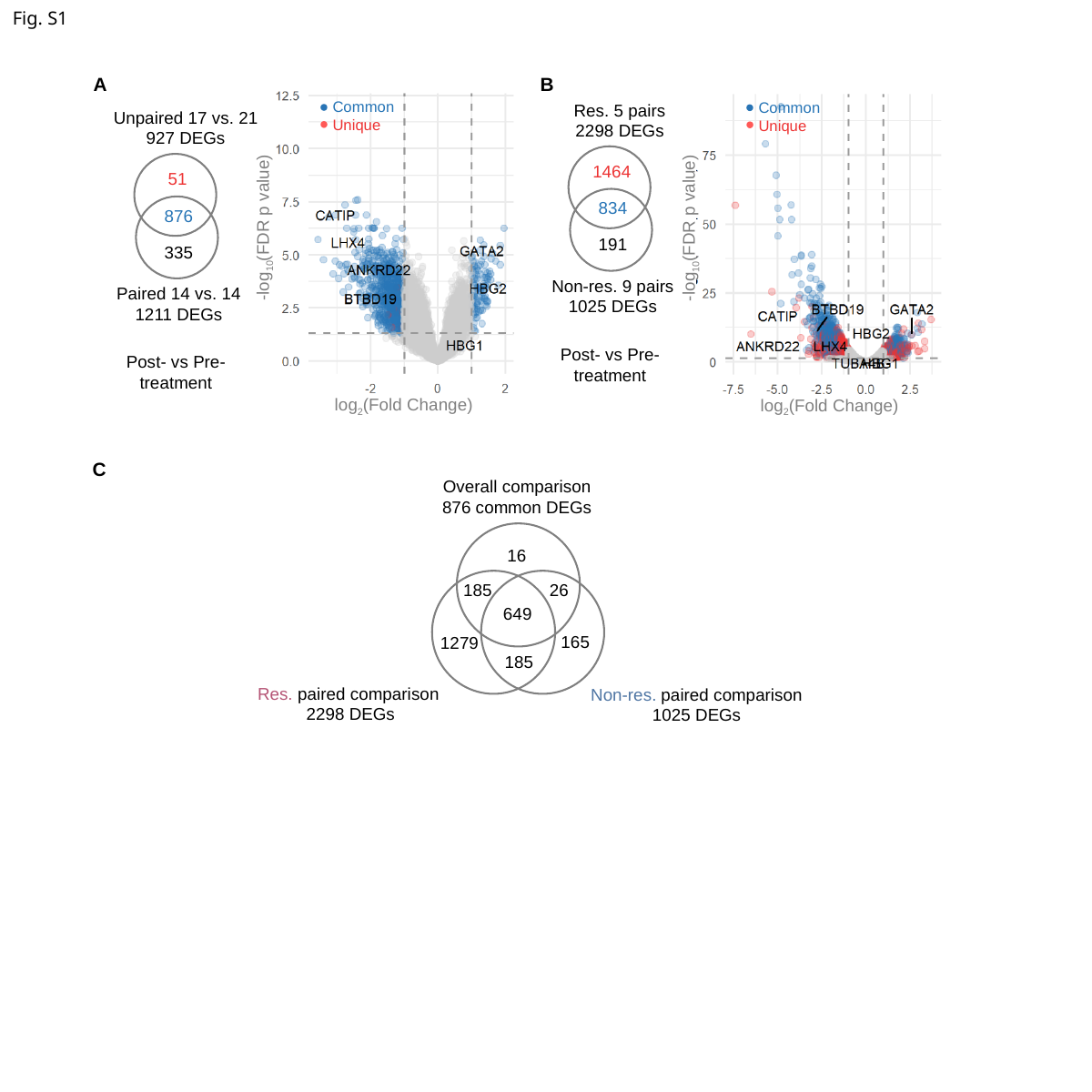

Fig. S1
A
B
Common
Unique
Common
Unique
Res. 5 pairs
2298 DEGs
Unpaired 17 vs. 21
927 DEGs
1464
51
834
876
-log_10_(FDR p value)
-log_10_(FDR p value)
191
335
Non-res. 9 pairs
1025 DEGs
Paired 14 vs. 14
1211 DEGs
Post- vs Pre-
treatment
Post- vs Pre-
treatment
log_2_(Fold Change)
log_2_(Fold Change)
C
Overall comparison
876 common DEGs
16
185
26
649
165
1279
185
Res. paired comparison
2298 DEGs
Non-res. paired comparison
1025 DEGs

### Slide 2
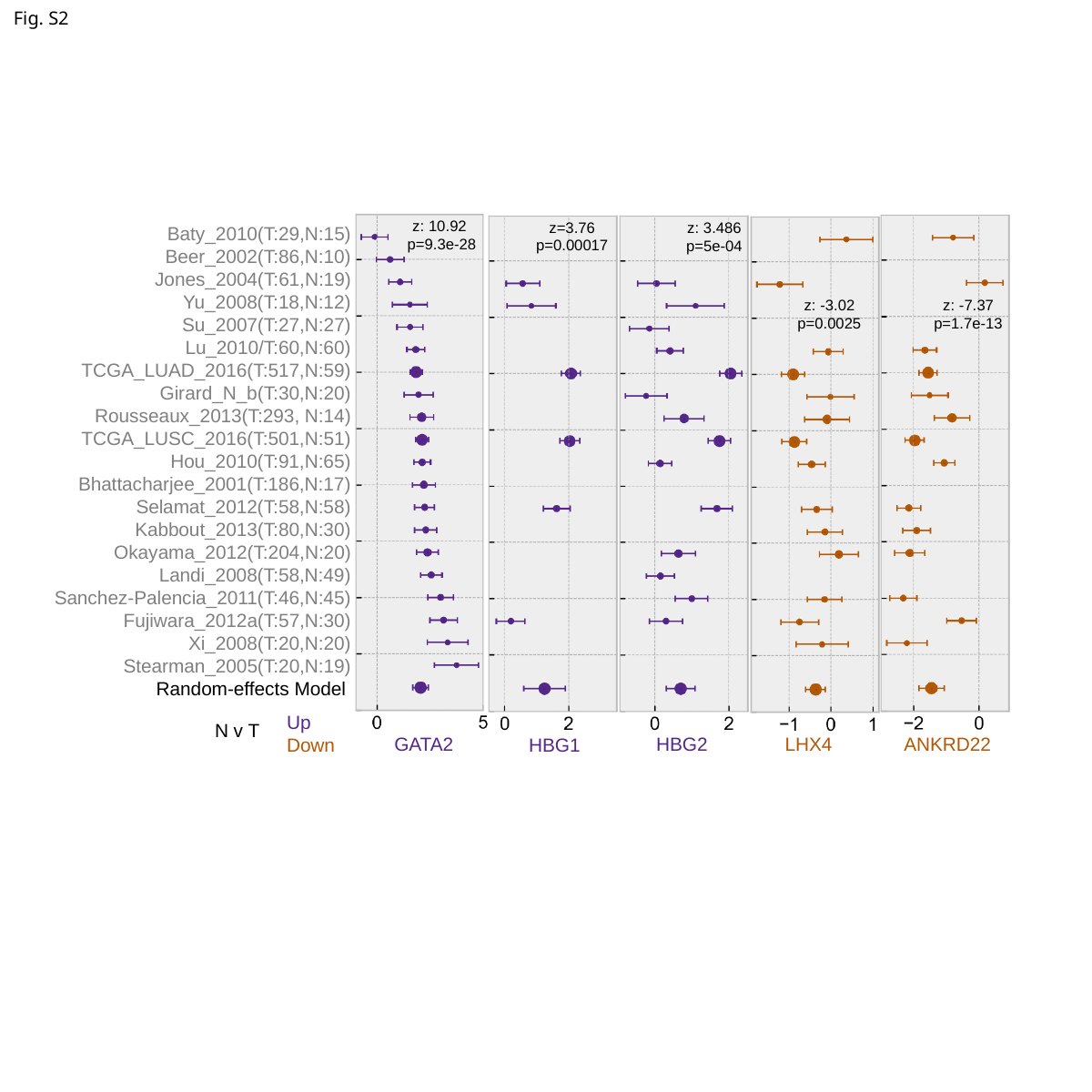

Fig. S2
z: 10.92
p=9.3e-28
z=3.76
p=0.00017
z: 3.486
p=5e-04
Baty_2010(T:29,N:15)
Beer_2002(T:86,N:10)
Jones_2004(T:61,N:19)
Yu_2008(T:18,N:12)
Su_2007(T:27,N:27)
Lu_2010/T:60,N:60)
TCGA_LUAD_2016(T:517,N:59)
Girard_N_b(T:30,N:20)
Rousseaux_2013(T:293, N:14)
TCGA_LUSC_2016(T:501,N:51)
Hou_2010(T:91,N:65)
Bhattacharjee_2001(T:186,N:17)
Selamat_2012(T:58,N:58)
Kabbout_2013(T:80,N:30)
Okayama_2012(T:204,N:20)
Landi_2008(T:58,N:49)
Sanchez-Palencia_2011(T:46,N:45)
Fujiwara_2012a(T:57,N:30)
Xi_2008(T:20,N:20)
Stearman_2005(T:20,N:19)
Random-effects Model
z: -3.02
p=0.0025
z: -7.37
p=1.7e-13
Up
Down
N v T
HBG2
LHX4
ANKRD22
GATA2
HBG1

### Slide 3
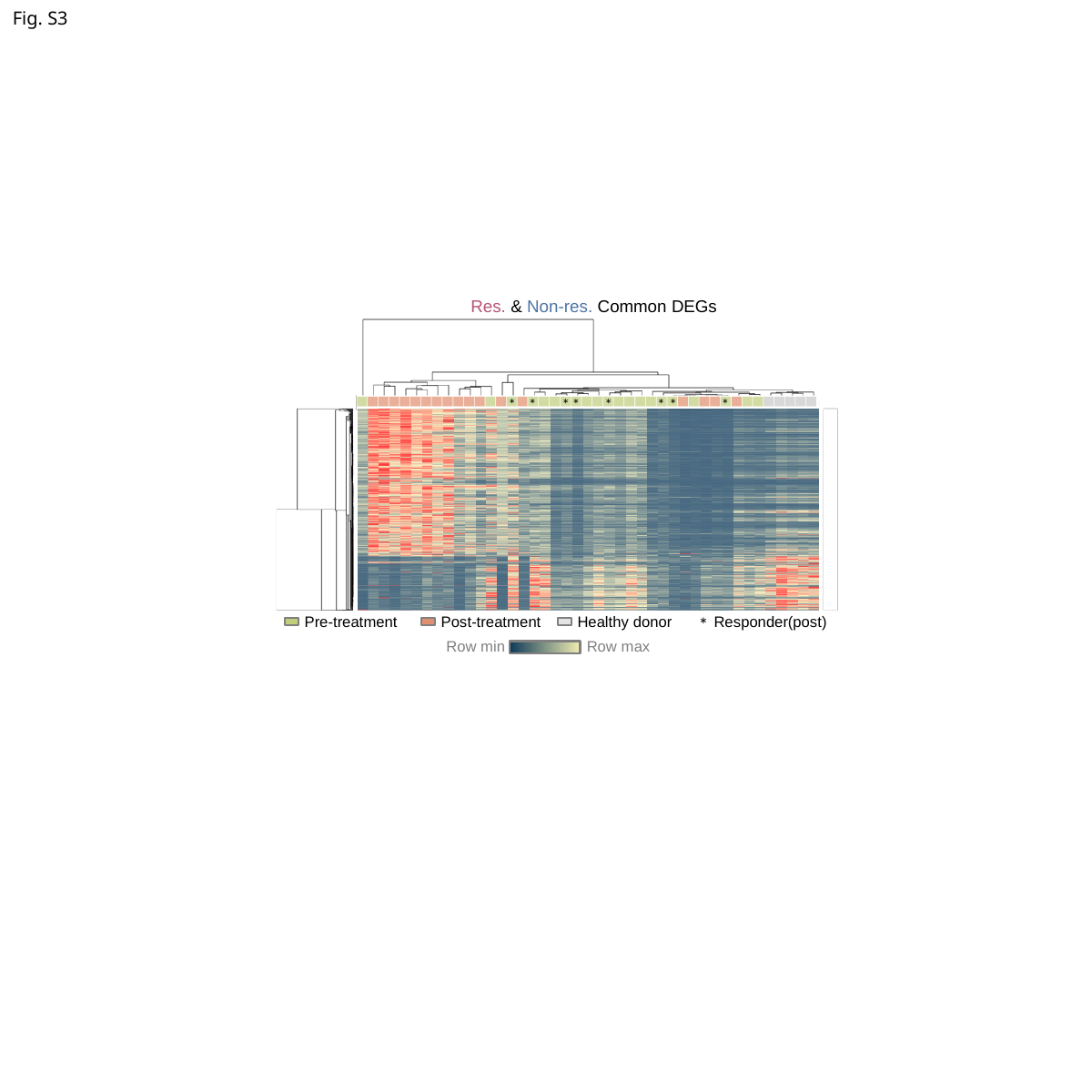

Fig. S3
Res. & Non-res. Common DEGs
*
*
*
*
*
*
*
*
*
Pre-treatment	Post-treatment	Healthy donor	Responder(post)
Row max
Row min

### Slide 4
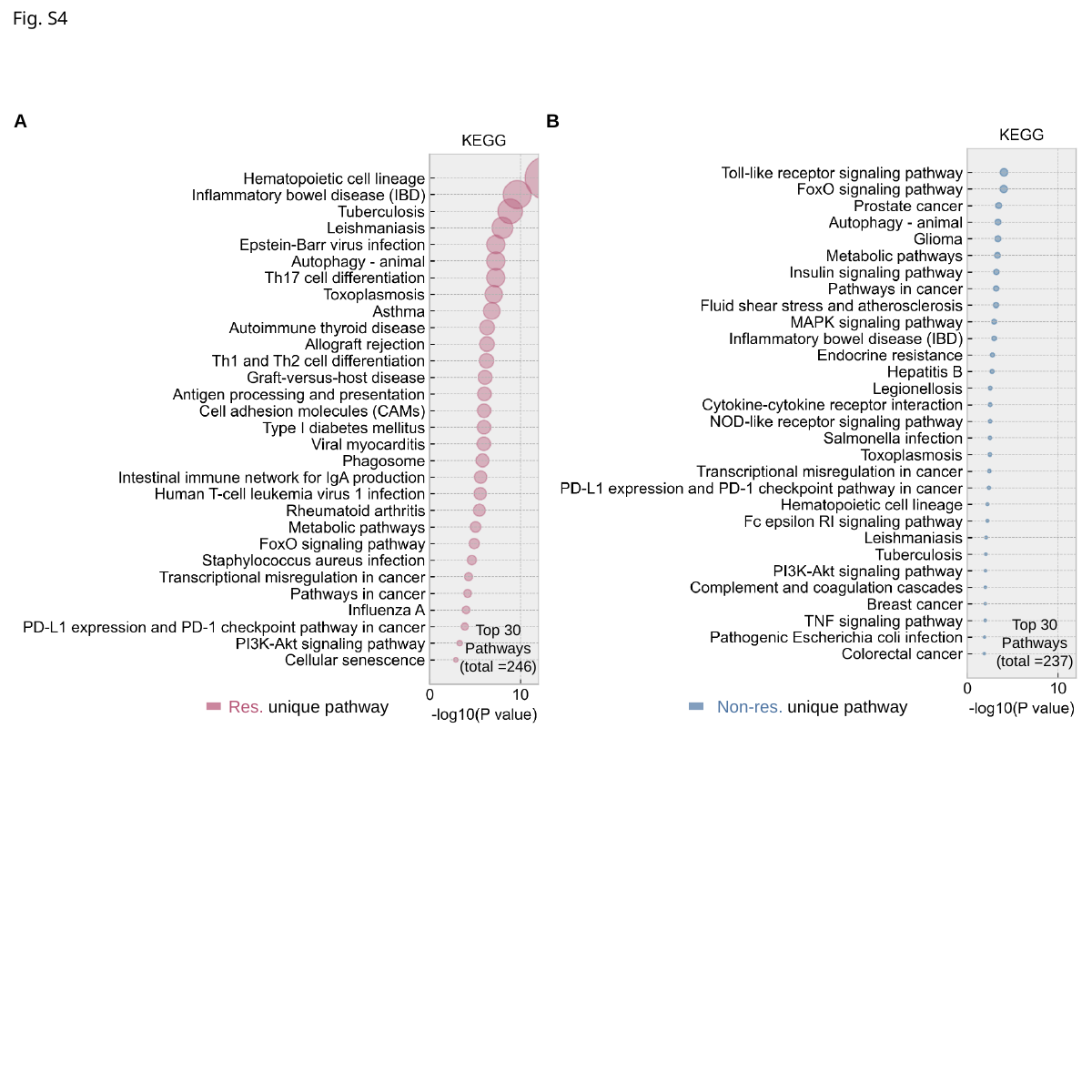

Fig. S4
A
B
Top 30 Pathways (total =237)
Top 30 Pathways (total =246)
Res. unique pathway
Non-res. unique pathway

### Slide 5
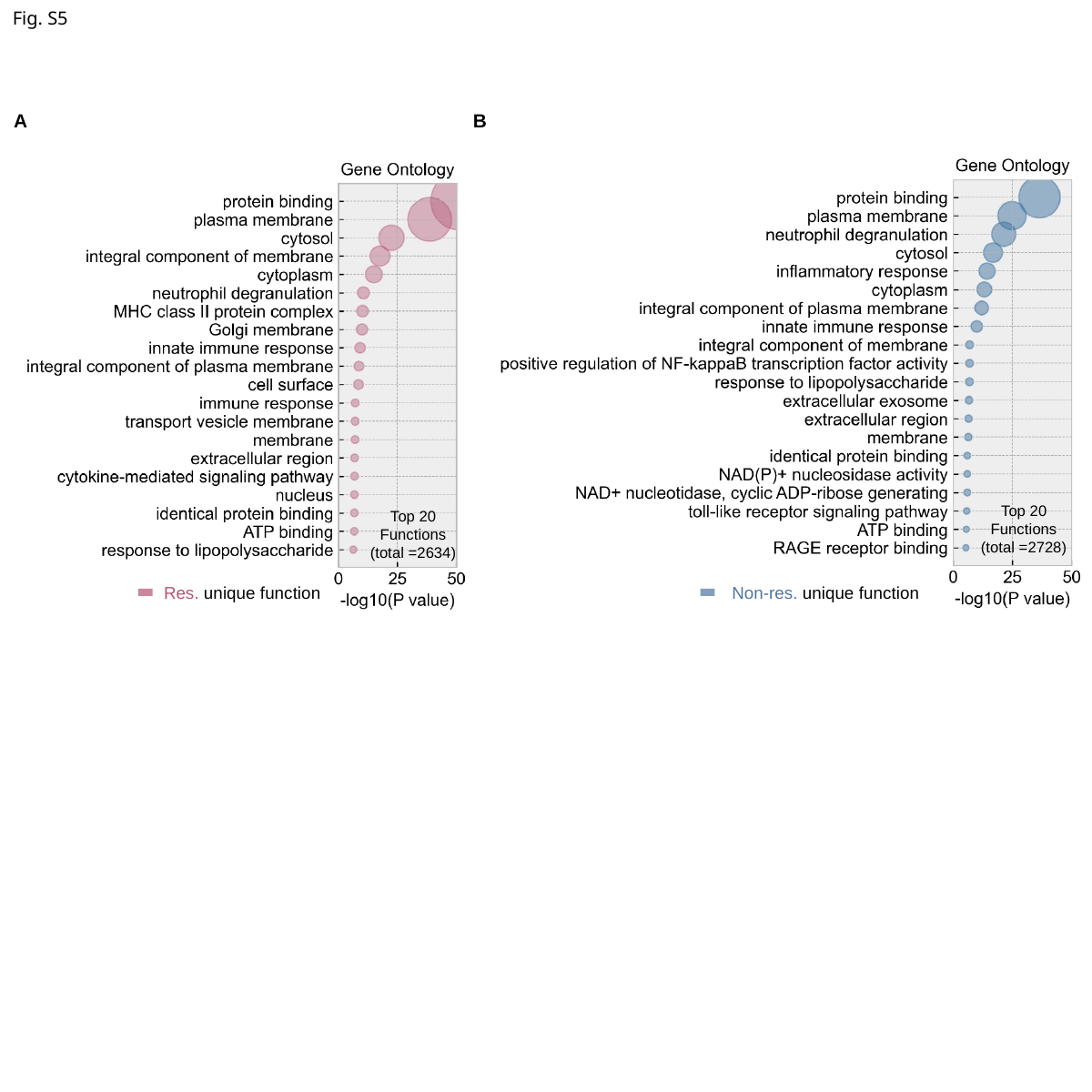

Fig. S5
A
B
Top 20 Functions (total =2728)
Top 20 Functions (total =2634)
Res. unique function
Non-res. unique function

### Slide 6
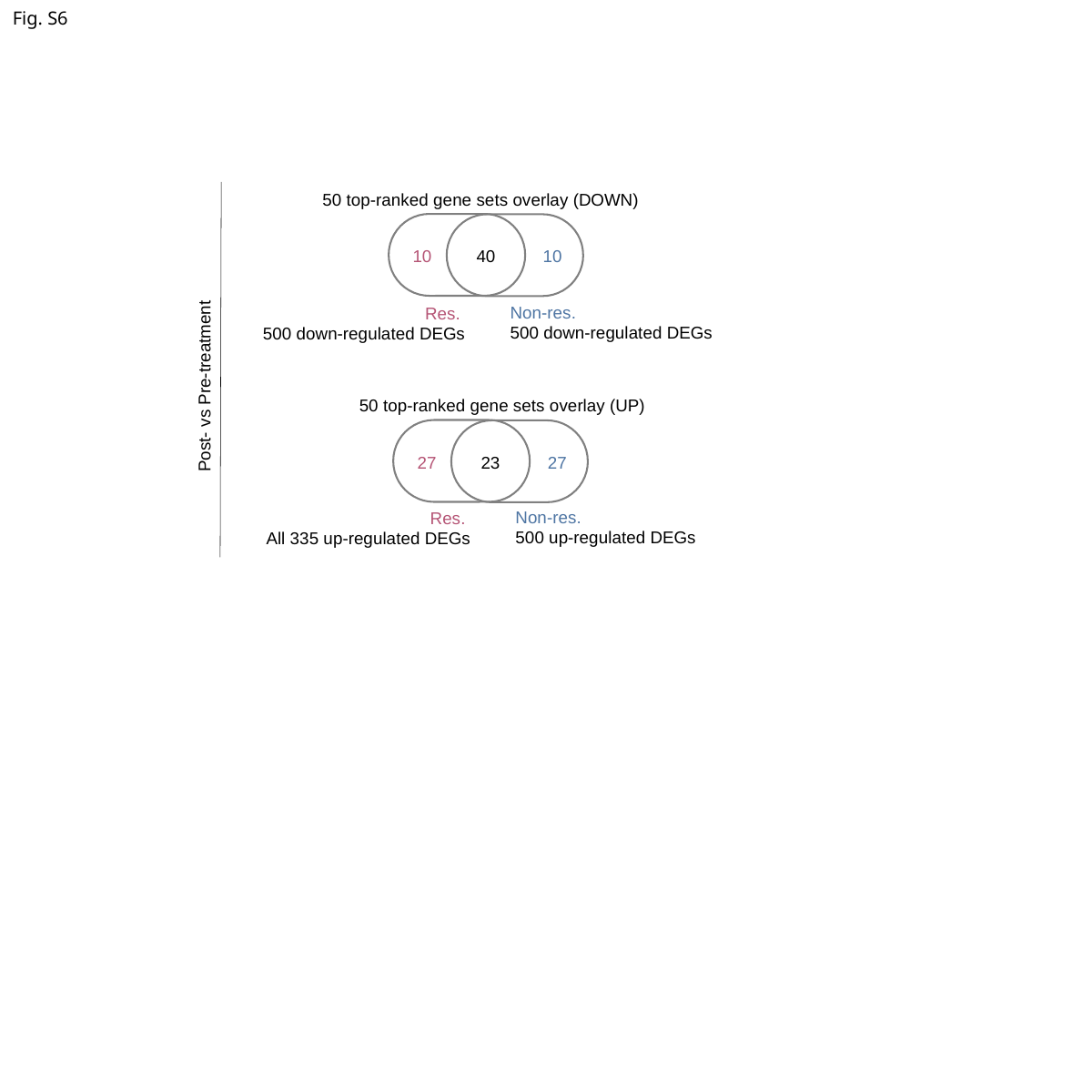

Fig. S6
50 top-ranked gene sets overlay (DOWN)
10
40
10
Non-res.
500 down-regulated DEGs
Res.
500 down-regulated DEGs
Post- vs Pre-treatment
50 top-ranked gene sets overlay (UP)
27
23
27
Non-res.
500 up-regulated DEGs
Res.
All 335 up-regulated DEGs

### Slide 7
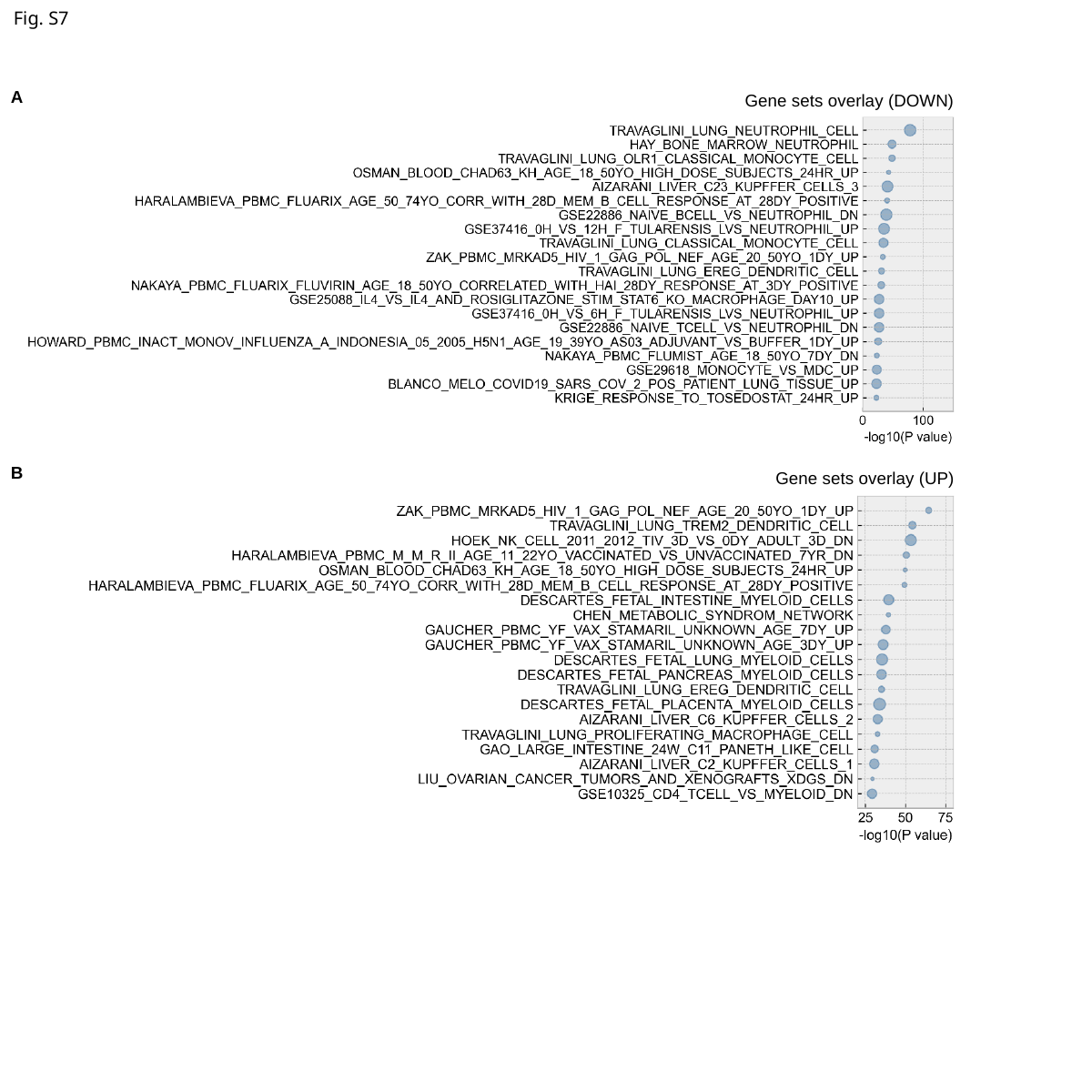

Fig. S7
A
Gene sets overlay (DOWN)
B
Gene sets overlay (UP)

### Slide 8
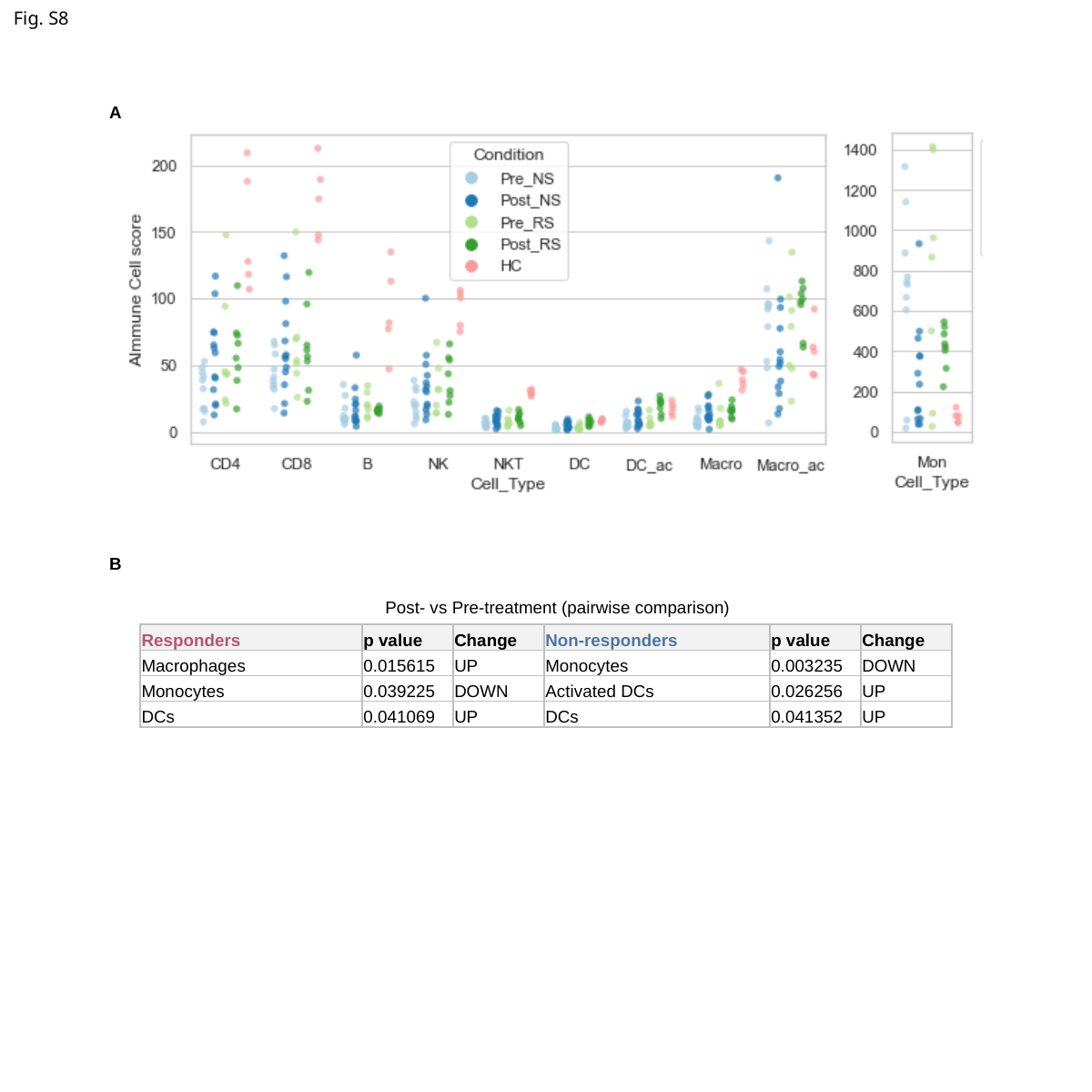

Fig. S8
A
B
Post- vs Pre-treatment (pairwise comparison)
| Responders | p value | Change | Non-responders | p value | Change |
| --- | --- | --- | --- | --- | --- |
| Macrophages | 0.015615 | UP | Monocytes | 0.003235 | DOWN |
| Monocytes | 0.039225 | DOWN | Activated DCs | 0.026256 | UP |
| DCs | 0.041069 | UP | DCs | 0.041352 | UP |

### Slide 9
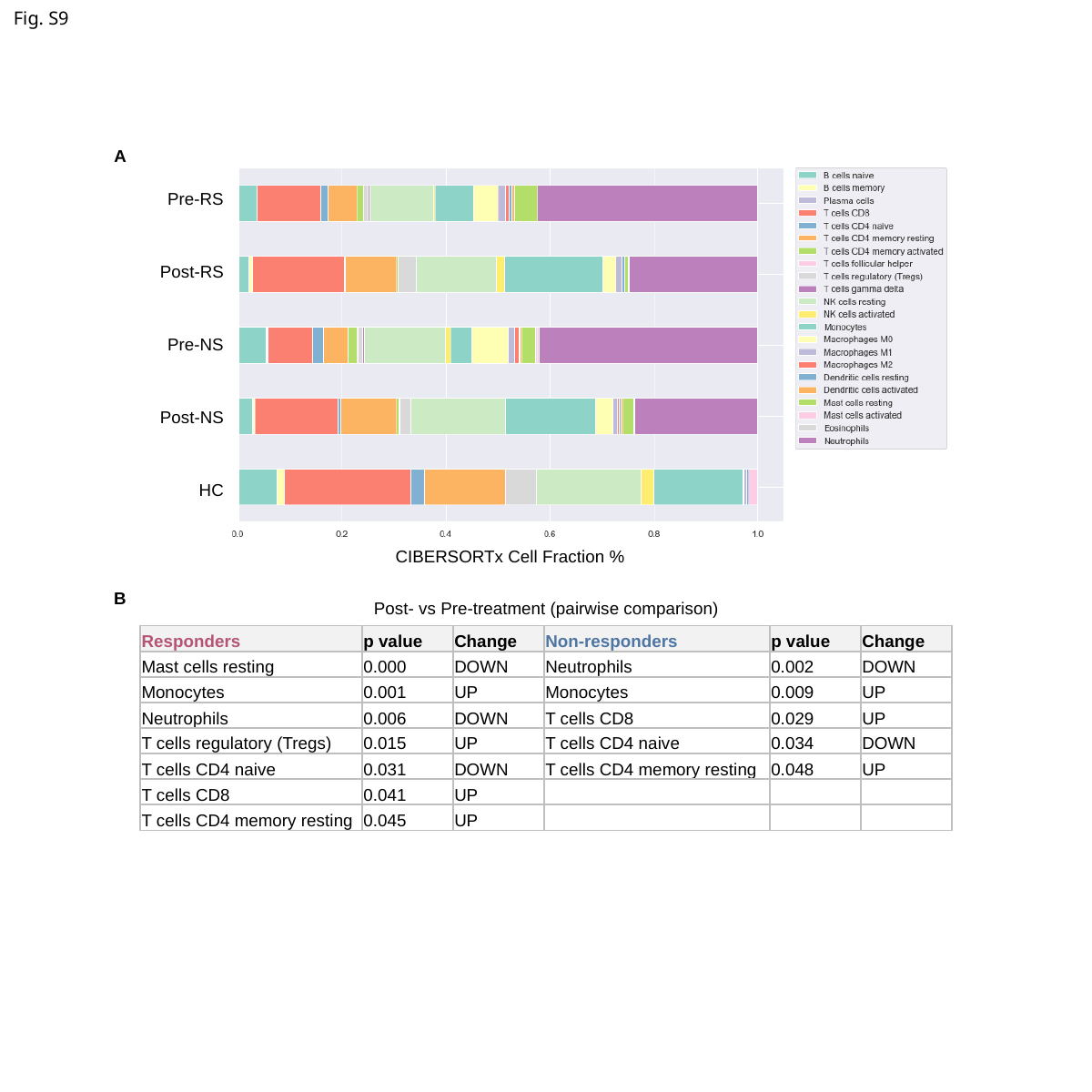

Fig. S9
A
Pre-RS
Post-RS
Pre-NS
Post-NS
HC
CIBERSORTx Cell Fraction %
B
Post- vs Pre-treatment (pairwise comparison)
| Responders | p value | Change | Non-responders | p value | Change |
| --- | --- | --- | --- | --- | --- |
| Mast cells resting | 0.000 | DOWN | Neutrophils | 0.002 | DOWN |
| Monocytes | 0.001 | UP | Monocytes | 0.009 | UP |
| Neutrophils | 0.006 | DOWN | T cells CD8 | 0.029 | UP |
| T cells regulatory (Tregs) | 0.015 | UP | T cells CD4 naive | 0.034 | DOWN |
| T cells CD4 naive | 0.031 | DOWN | T cells CD4 memory resting | 0.048 | UP |
| T cells CD8 | 0.041 | UP | | | |
| T cells CD4 memory resting | 0.045 | UP | | | |

### Slide 10
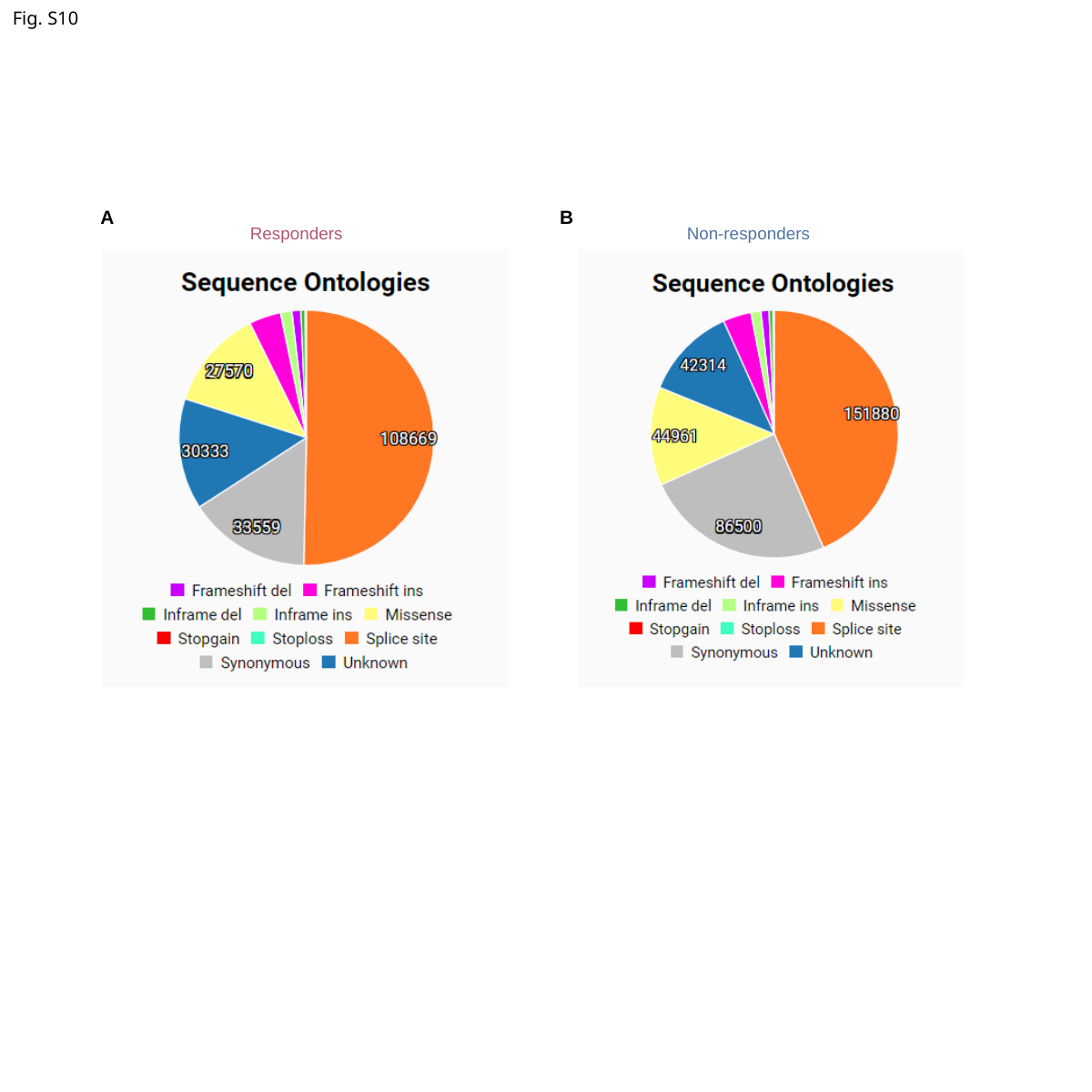

Fig. S10
A
B
Responders
Non-responders

### Slide 11
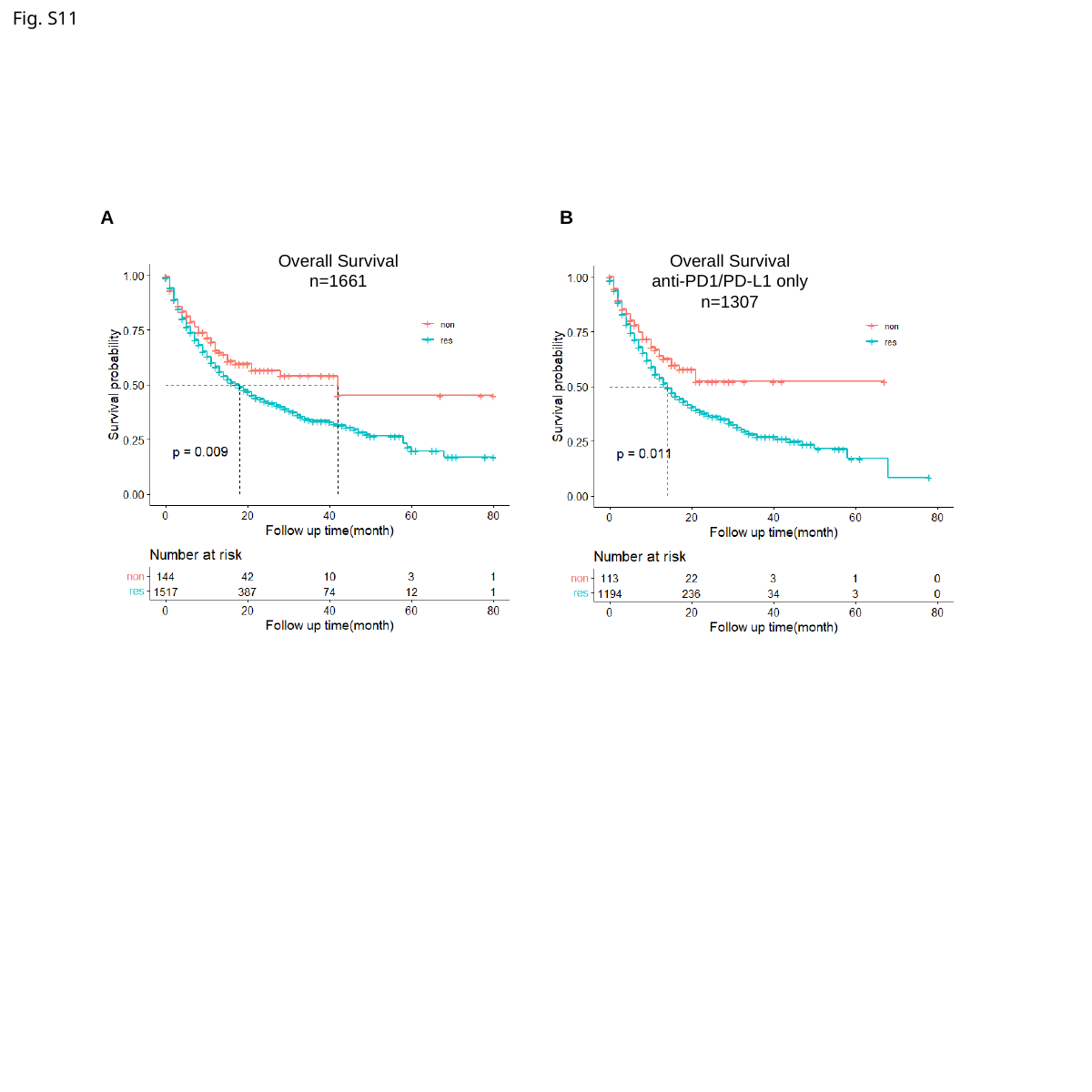

Fig. S11
A
B
Overall Survival
n=1661
Overall Survival
anti-PD1/PD-L1 only
n=1307
