## Supplemental Figure Legends for "Blood-based Genomic and Cellular Determinants of Response to Neoadjuvant PD-1 Blockade in Patients with Non-Small-Cell Lung Cancer"

**Supplementary documents**

**Supplementary Figure S1.** A) Venn diagram and volcano plot of overall DEGs in unpaired and paired comparisons of post- versus pre-treatment blood samples. Unique DEGs identified from unpaired comparison and Common DEGs identified from both pairwise and unpaired comparisons are color-coded as annotated. B) Venn diagram and volcano plot of DEGs identified in comparison of post- versus pre-treatment blood samples from non-responders and responders. DEGs only seen in responders (Unique) and DEGs seen in both non-responders and responders (Common) are highlighted. C) Venn diagram of DEGs identified in comparisons of post- versus pre-treatment blood samples. These DEGs were identified respectively from responders (paired comparison), non-responders (paired comparison), and from 876 common DEGs in overall comparison.

**Supplementary Figure S2.** Forest plots showing the standardized mean of gene expression difference between normal and tumor tissue as estimated from multiple studies (collected from LCE database). The leftmost column shows the included studies by the first author’s name and publication year and followed by the cohort size. The circles lined up in each column represent the effect estimates from individual studies and the very bottom circles show the pooled result for each gene as annotated. The size of each circle indicates the cohort size of individual study. The horizontal lines through the boxes illustrate the length of the 95% confidence interval in both positive and negative sides. Random-effects model was utilized to evaluate the overall effect as described by z-score and p value. N, normal lung tissue; T, lung cancer tissue; v, versus.

**Supplementary Figure S3.** The hierarchical clustering of all study samples according to their profiles of Common DEGs identified from both responder and non-responder. The heatmap visualized the relative expression level of each DEG. Pre- and post-treatment status and responder labels are color-coded and annotated.

**Supplementary Figure S4.** Bubble plots of the top 30 KEGG pathways regulated in responders (A) and non-responders (B). Bubble with bigger size stands for smaller p value and higher significance. Res, responders; Non-res, non-responders.

**Supplementary Figure S5.** Bubble plots of the top 20 unique GO items regulated in responders (A) and non-responders (B). Bubble with bigger size stands for smaller p value and higher significance. Res, responders; Non-res, non-responders.

**Supplementary Figure S6.** Venn diagram of the top 50 gene sets downregulated (upper) or downregulated (lower) identified in comparison of post- versus pre-treatment samples. The count of DEGs used to carry out the GSEA were provided as annotated. The number of unique gene sets are colored in red (responder) or blue (non-responder) while the shared gene sets are annotated in grey text. Res, responders; Non-res, non-responders.

**Supplementary Figure S7.** Bubble plots of top 20 gene sets down-regulated (A) and up-regulated (B) in non-responders. Bubble with bigger size stands for higher k/K value ratio and larger fraction of gene was matched with a certain reference gene set.

**Supplementary Figure S8.** Immune cell abundance scores calculated by AImmune. A) Dot plot showing AImmune cell abundance scores of 10 immune cell subsets across five study groups as color-coded and annotated. B) Immune cell subsets with AImmune scores that are significantly (p < 0.05) different across post- vs pre-treatment samples. Pre-NS, pre-treatment samples from non-responders; Post-NS, post-treatment samples from non-responders; Pre-RS, pre-treatment samples from responders; Post-RS, post-treatment from responders; HC, healthy control; CD4, CD4+ T cells; CD8, CD8+ T cells; B, B cells; NK, natural killer cells; NKT, natural killer T cells; DC, dendritic cells; DC_ac, activated dendritic cells; Macro, macrophages; Macro_ac, activated macrophages; Mon, monocytes. All p values were calculated via pairwise comparisons.

**Supplementary Figure S9.** Immune cell fractions estimated by CIBERSORTx. A) Stacked bar plot showing individual fractions of 22 immune cell subsets in five study groups color-coded and annotated. The different conditions are shown in different colors. B) Immune cell subsets with CIBERSORTx fractions that are significantly (p < 0.05) different across post- vs pre-treatment samples. Pre-NS, pre-treatment samples from non-responders; Post-NS, post-treatment samples from non-responders; Pre-RS, pre-treatment samples from responders; Post-RS, post-treatment from responders; HC, healthy control. All p values were calculated via pairwise comparisons.

**Supplementary Figure S10.** Pie charts showing distribution and counts of the reported mutations grouped by sequence ontology as identified in responders (A) and non-responders (B).

**Supplementary Figure S11.** The Kaplan Meier overall survival curves of two patient groups defined by 4 non-responder markers combined as one panel. Curves were generated for all immunotherapy patients (A) and patients received anti-PD1/PD-L1 therapy only (B).

**Table S1** DEG lists identified from this study

**Table S1a** List of DEGs identified in unpaired comparison (all subjects), n=927

**Table S1b** List of DEGs identified in paired comparison (all subjects), n=1211

**Table S1c** List of DEGs identified in responders, n=2298

**Table S1d** List of DEGs identified in non-responders, n=1025

**Table S2** List of PCR primers used in this study
